# CLINPREAI: AN AGENTIC AI SYSTEM FOR EARLY POSTPARTUM DEPRESSION RISK PREDICTION FROM MULTIMODAL EHR DATA

**DOI:** 10.1101/2025.11.14.25340265

**Authors:** Daniel Palacios, Sukru Aras, Yi Zhong, Jeff Zhao, Sasidhar Pasupuleti, Hyun-Hwan Jeong, Emily S. Miller, Terri L. Fletcher, Alison N. Goulding, Hu Chen, Zhandong Liu

## Abstract

Postpartum depression (PPD) affects 10–15% of individuals annually, yet early identification and treatment remains challenging. We introduce ClinPreAI, a novel agentic AI system that autonomously designs, implements, and evaluates machine learning solutions for PPD risk prediction using multimodal electronic health record data. We analyzed data from 4,161 pregnant individuals hospitalized prior to delivery for medical or obstetrical complications at Texas Children’s Hospital (2012–2025), extracting 27 structured clinical variables and social worker notes. The primary outcome was Edinburgh Postnatal Depression Scale (EPDS) score ≥10 (31.0% prevalence) within 6 months after delivery, indicating clinically significant depressive symptoms. ClinPreAI operates through five specialized modules that iteratively refine predictive models through autonomous experimentation. ClinPreAI demonstrated strong performance across modalities. On structured data, it achieved F1: 0.68 ± 0.03, outperforming traditional AutoML (F1: 0.64 ± 0.02) and commercial solutions (AWS Canvas F1: 0.54–0.55). On multimodal data, ClinPreAI achieved F1: 0.65 ± 0.04, matching custom LLM-XGBoost (F1: 0.65 ± 0.01) and outperforming zero-shot models (Claude Opus F1: 0.51–0.52). This represents the first application of agentic AI to perinatal mental health prediction. Our results demonstrate that autonomous AI agents can democratize sophisticated predictive modeling in clinical settings, which is particularly valuable where domain experts lack ML training. By automating experimentation and debugging, agentic systems lower barriers to developing robust clinical prediction tools while maintaining interpretability.

## Introduction

Postpartum depression (PPD) is common, affecting 10–15% of individuals annually, with symptoms persisting beyond six months in 25–50% of affected people [1]. Mental health conditions, such as PPD, are the leading preventable cause of maternal mortality in the United States [2]. Meta-analytic studies identify key predictors including history of depression prior to pregnancy, antenatal depression and anxiety, major stressful life events, and inadequate social support [3, 4, 5, 6, 7, 8, 9, 10]. Hospitalized antepartum patients, defined as women hospitalized for medical or obstetric complications prior to delivery, are at particularly elevated risk for postpartum mental health conditions. Approximately one in three patients screens positive for depression during their antepartum hospitalization [11], which is twice the prevalence in the general obstetric population; these patients are also at increased risk for postpartum depression [12]. This high-risk population has specific mental health risk factors, including pregnancy complications, hospitalization-related isolation, and increased likelihood of preterm birth and subsequent neonatal intensive care unit admission [11, 13, 14, 15]. The Edinburgh Postnatal Depression Scale (EPDS) [16] remains the primary screening tool for perinatal depression, with universal screening recommended twice during pregnancy and again during the postpartum period [17]. Unfortunately, our current approach to perinatal mental health screening and treatment is inadequate, with a large meta-analysis showing that the minority of patients (∼ 40%) receive recommended depression screening and the small minority (∼ 7%) receive adequate treatment [18]. Additionally, the EPDS suffers from scoring errors in up to 17% of cases [19]. Developing more accurate approaches to identify those at highest risk of PPD would enable timely intervention and direction of limited mental health resources [20, 21, 22, 23] to those most in need. The aim of this study was to utilize multimodal electronic health record (EHR) data from a single antepartum hospitalization to predict PPD. Because predictions based on a single antepartum hospitalization reflect the information available at that time, this study evaluates whether meaningful PPD risk stratification is feasible early in pregnancy care.

Traditional machine learning approaches for PPD prediction have employed logistic regression, random forests, and gradient boosting methods [24, 25, 26], with some neural network implementations using standardized databases [27]. However, most models utilize only structured EHR variables, omitting unstructured clinical narratives, such as notes by social workers that document important psychosocial stressors rarely captured in coded fields [28]. This contrasts with predictive modeling in other clinical domains where multimodal EHR+text models outperform structured-only approaches [29, 30, 31, 32]. Moreover, published PPD models typically lack provisions for dynamic adaptation or human-in-the-loop refinement [33], limiting real-world deployment.

The emergence of agentic AI systems leveraging large language models (LLMs) for autonomous reasoning, planning, and task execution [34] offers transformative potential for clinical predictive modeling. Unlike traditional ML pipelines requiring manual feature engineering and model selection, AI agents autonomously design end-to-end solutions through iterative experimentation, providing: (1) systematic solution space exploration via autonomous planning; (2) self-verification with output validation and error correction; (3) iterative refinement through feedback loops; and (4) interpretability through explicit reasoning traces—critical for clinical deployment. Recent healthcare applications include disease mechanism discovery [35], biomedical terminology normalization [36], research workflow automation [37], and multi-specialty diagnostic consensus [38, 39, 40]. Machine learning-focused frameworks demonstrate training-free experiment automation [41], self-evolving architectures with cloud platform support [42], and data science workflow optimization [43]. However, as shown in Table 1, existing frameworks vary considerably in capabilities, with none specifically designed for clinical predictive modeling.

**Table 1:** Comparison of AutoML and Agentic AI Methods.

| Method | Planning | Verification | Full Pipeline | Task Agnostic | Training Free | Multimodality | Error Analysis | Interpretation | AWS Bedrock Support | Dynamic Model Training |
| --- | --- | --- | --- | --- | --- | --- | --- | --- | --- | --- |
| ClinPreAI AutoML |  |  | ✓ | ✓ |  | ✓ | ✓ |  | ✓ |  |
| ClinPreAI Agent | ✓ | ✓ | ✓ | ✓ |  | ✓ |  | ✓ | ✓ | ✓ |
| AWS Canvas |  |  |  | ✓ |  |  |  |  |  |  |
| AutoML-Agent [41] | ✓ | ✓ | ✓ | ✓ | ✓ | ✓ |  |  |  | ✓ |
| SELA [42] | ✓ | ✓ |  | ✓ |  | ✓ |  |  |  | ✓ |
| DS-Agent [43] | ✓ | ✓ |  | ✓ |  | ✓ |  |  |  |  |

Traditional AutoML platforms democratize machine learning [44] but remain constrained by predefined pipelines unable to reason about domain-specific requirements. Clinical frameworks like EHR-ML streamline data preprocessing [45] yet require substantial human oversight for critical decisions. Integrating agentic capabilities into clinical ML workflows thus addresses longstanding challenges in model development, validation, and deployment.

This study introduces ClinPreAI, a novel agentic AI system for autonomous development and refinement of PPD risk prediction models using multimodal EHR data. We analyze data from 4,161 pregnant individuals hospitalized prior to delivery for medical or obstetrical complications between 2012 and 2025 at Texas Children’s Hospital, a regional referral center. We integrated 27 structured clinical variables with unstructured social worker notes from first-admission encounters. Social worker notes often capture psychosocial risk factors that don’t appear in structured fields (or appear late or incompletely), including housing instability, food insecurity, intimate partner violence (IPV), social support, transportation barriers, substance use context, prior mental health history, custody concerns, immigration/legal stressors, and caregiver strain [46]. The primary outcome is postpartum EPDS score ≥ 10 (31.0% prevalence), indicating the presence of clinically significant PPD symptoms. ClinPreAI operates through five specialized modules—research, planning, coding, debugging, and interpretation—handling the complete ML pipeline from preprocessing through results interpretation. Our key contributions include:

- **Agentic multimodal framework**. Purpose-built system for clinical prediction achieving F1 = 0.68 *±* 0.03, outperforming traditional AutoML (F1 = 0.64 *±* 0.02), commercial platforms (F1 = 0.54–0.55), and LLM prompting (F1 = 0.51–0.52), matching custom LLM + XGBoost model (F1 = 0.65 *±* 0.01).
- **Comprehensive benchmarking**. Systematic comparison across traditional ML, AutoML workflows, ML commercial platforms, LLM-based approaches, and agentic systems, establishing deployment trade-offs.
- **Enhanced interpretability**. Ablation studies, error analysis, and unsupervised clustering reveal clinical insights including dominance of prior mental health history as risk factor.
- **Generalizable clinical AI template**. Reusable agent architecture for multimodal clinical outcomes prediction tasks.
- **Single-encounter prediction**. Unlike models trained at delivery or using longitudinal data [47], we study early risk stratification from first antepartum hospitalization.

This work establishes a methodological foundation for autonomous AI systems in clinical decision support, advancing both PPD prediction capabilities and broader applications of agentic AI in healthcare.

## Results

### Prior Anxiety and Depression Diagnoses, Psychotropic Medication Use, and Social Work Service Utilization Associate with High Postpartum EPDS Scores

Of the 4,161 patients included in the analysis, 2,870 (69.0%) had postpartum EPDS scores below 10, while 1,291 (31.0%) had scores of 10 or above, indicating elevated postpartum depression (PPD) symptoms—a commonly applied screening threshold for possible/minor depression [48]. Table 2 presents the demographic, clinical, and social characteristics of the cohort stratified by EPDS score. We modeled EPDS as a binary outcome (EPDS ≥ 10 vs. *<* 10) to align with clinically interpretable screening decisions (screen-positive vs. screen-negative) and to facilitate evaluation using actionable classification metrics. This approach also avoids imposing a linear relationship across the full EPDS range, which may not hold uniformly for symptom changes at different score levels.

**Table 2:** Patient characteristics stratified by Edinburgh Postnatal Depression Scale (EPDS) score.

| Characteristic | Overall<br>(N = 4161) | EPDS < 10<br>(n = 2870, 69.0%) | EPDS ≥ 10<br>(n = 1291, 31.0%) | P-value<sup>a</sup> |
| --- | --- | --- | --- | --- |
| **Demographics** |  |  |  |  |
| **Maternal age (years)** |  |  |  |  |
| Median (IQR) | 30.6 (25.9–34.9) | 30.6 (26.0–35.0) | 30.6 (25.8–34.8) | 0.513 |
| **Gestational age on admission (weeks)** |  |  |  |  |
| Median (IQR) | 29.7 (24.7–33.3) | 29.9 (25.0–33.4) | 29.0 (24.0–33.3) | 0.030 |
| **Length of stay (days)** |  |  |  |  |
| Median (IQR) | 6.0 (3.0–9.0) | 6.0 (3.0–9.0) | 5.0 (3.0–9.0) | 0.578 |
| **Adolescent maternal age (< 20 years), n (%)** | 193 (4.6) | 127 (4.4) | 66 (5.1) | 0.445 |
| **Gravidity > 1, n (%)** | 1670 (40.1) | 1200 (41.8) | 470 (36.4) | 0.003 |
| **Gestational age category, n (%)** |  |  |  | 0.046 |
| < 28 weeks | 1675 (40.3) | 1117 (38.9) | 558 (43.2) |  |
| 28–32 weeks | 1000 (24.0) | 698 (24.3) | 302 (23.4) |  |
| > 32 weeks | 1486 (35.7) | 1055 (36.8) | 431 (33.4) |  |
| **Race, n (%)** |  |  |  | 0.054 |
| White | 2713 (65.2) | 1898 (66.1) | 815 (63.1) |  |
| Black | 1164 (28.0) | 766 (26.7) | 398 (30.8) |  |
| Asian | 219 (5.3) | 157 (5.5) | 62 (4.8) |  |
| Other | 65 (1.6) | 49 (1.7) | 16 (1.2) |  |
| **Hispanic or Latino ethnicity, n (%)** | 1445 (34.7) | 1046 (36.4) | 399 (30.9) | 0.003 |
| **Spanish language, n (%)** | 266 (6.4) | 207 (7.2) | 59 (4.6) | 0.004 |
| **Marital status, n (%)** |  |  |  | 0.003 |
| Married | 2365 (56.8) | 1692 (59.0) | 673 (52.1) |  |
| Single | 1468 (35.3) | 967 (33.7) | 501 (38.8) |  |
| Other | 328 (7.9) | 211 (7.4) | 117 (9.1) |  |
| **Insurance type, n (%)** |  |  |  | 0.004 |
| Private | 2300 (55.3) | 1636 (57.0) | 664 (51.4) |  |
| Government | 1797 (43.2) | 1187 (41.4) | 610 (47.3) |  |
| Self-pay | 64 (1.5) | 47 (1.6) | 17 (1.3) |  |
| **Medical History** |  |  |  |  |
| **Any history of mental illness, n (%)** | 1451 (34.9) | 753 (26.2) | 698 (54.1) | 0.003 |
| **Pregestational diabetes, n (%)** | 425 (10.2) | 263 (9.2) | 162 (12.5) | 0.003 |
| **Chronic hypertension, n (%)** | 632 (15.2) | 402 (14.0) | 230 (17.8) | 0.004 |
| **Chronic medical conditions, n (%)** | 2402 (57.7) | 1582 (55.1) | 820 (63.5) | 0.003 |
| **Pregnancy Complications** |  |  |  |  |
| **Premature rupture of membranes, n (%)** | 581 (14.0) | 389 (13.6) | 192 (14.9) | 0.391 |
| **Preterm labor, n (%)** | 122 (2.9) | 77 (2.7) | 45 (3.5) | 0.281 |
| **Placenta previa, n (%)** | 209 (5.0) | 146 (5.1) | 63 (4.9) | 0.837 |
| **Placenta accreta spectrum, n (%)** | 153 (3.7) | 100 (3.5) | 53 (4.1) | 0.445 |
| **Gestational diabetes, n (%)** | 333 (8.0) | 226 (7.9) | 107 (8.3) | 0.724 |
| **Multiple gestation, n (%)** | 442 (10.6) | 315 (11.0) | 127 (9.8) | 0.393 |
| **Social Worker Notes** |  |  |  |  |
| **Social worker notes present, n (%)** | 1900 (45.7) | 1235 (43.0) | 665 (51.5) | <0.001 |
| **Words in notes per patient** |  |  |  |  |
| Median (IQR) | 707.5 (354.8–1062.5) | 694.0 (333.5–1013.5) | 753.0 (379.0–1162.0) | 0.003 |
| **HPO codes per patient** |  |  |  |  |
| Median (IQR) | 5.0 (3.0–8.0) | 4.0 (3.0–6.0) | 6.0 (3.0–10.0) | <0.001 |

<sup>a</sup>Adjusted P-values are reported. Continuous variables are summarized as median (IQR). EPDS, Edinburgh Postnatal Depression Scale; IQR, interquartile range; HPO, Human Phenotype Ontology.

**Table 3:**
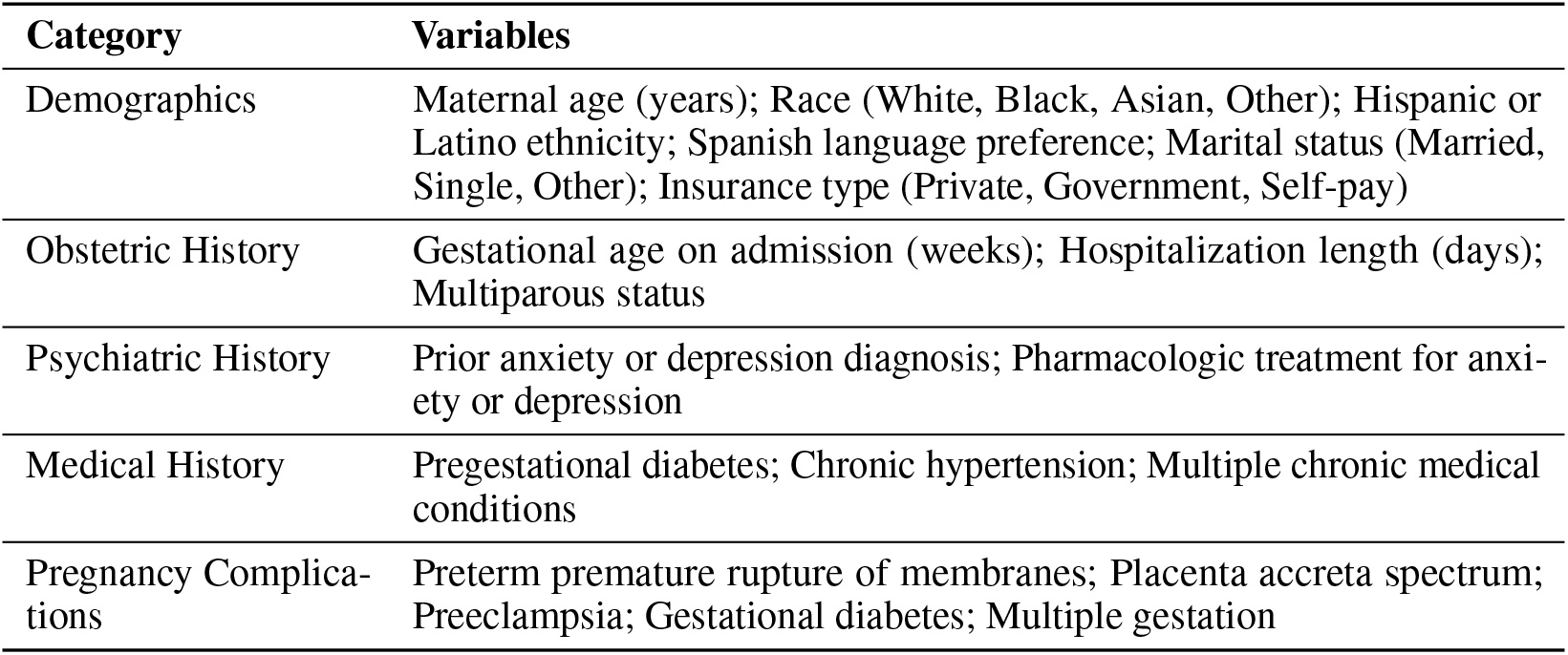
Structured variables extracted from the electronic health record (EHR)

| Category | Variables |
| --- | --- |
| Demographics | Maternal age (years); Race (White, Black, Asian, Other); Hispanic or Latino ethnicity; Spanish language preference; Marital status (Married, Single, Other); Insurance type (Private, Government, Self-pay) |
| Obstetric History | Gestational age on admission (weeks); Hospitalization length (days); Multiparous status |
| Psychiatric History | Prior anxiety or depression diagnosis; Pharmacologic treatment for anxiety or depression |
| Medical History | Pregestational diabetes; Chronic hypertension; Multiple chronic medical conditions |
| Pregnancy Complications | Preterm premature rupture of membranes; Placenta accreta spectrum; Preeclampsia; Gestational diabetes; Multiple gestation |

Several sociodemographic factors were associated with elevated depressive symptoms after adjustment for multiple testing. Patients with EPDS ≥ 10 were less likely to be married (52.1% vs. 59.0%, p=0.003), more likely to have government insurance (47.3% vs. 41.4%, p=0.004), and less likely to be of Hispanic/Latino ethnicity (30.9% vs. 36.4%, p=0.003) or Spanish-speaking (4.6% vs. 7.2%, p=0.004). Patients with elevated scores were admitted at slightly earlier gestational ages (mean 28.2 vs. 28.8 weeks, p=0.030) and were more likely to be admitted before 28 weeks gestation (43.2% vs. 38.9%, p=0.046).

The largest difference was observed in mental health history: 54.1% of patients with elevated postpartum EPDS scores had a documented history of mental illness, compared to 26.2% in the lower EPDS group (p<0.003). Patients with postpartum EPDS ≥ 10 also had higher rates of pregestational diabetes (12.5% vs. 9.2%, p=0.003), chronic hypertension (17.8% vs. 14.0%, p=0.004), and chronic medical conditions overall (63.5% vs. 55.1%, p=0.003). No significant differences were observed in maternal age, length of stay, or most pregnancy complications between the two groups.

Social work documentation patterns differed between groups. Overall, 1,900 patients (45.7% of the cohort) had social worker notes present. Among these, patients with elevated postpartum EPDS scores were more likely to have social worker notes documented compared to those with lower scores (51.5% vs. 43.0%, p<0.001). Among patients with social worker notes, those with a postpartum EPDS ≥ 10 had more extensive documentation (median 753 vs. 694 words per note, p=0.003) and more Human Phenotype Ontology (HPO) codes assigned (median 6 vs. 4 codes, p<0.001), suggesting greater psychosocial complexity or need for services. HPO codes example for this context includes: “Depressed mood” (HP:0000716).

These statistical differences between the groups raised an important question: do patients with elevated postpartum EPDS scores represent a distinct, identifiable subgroup, or do they exist along a continuum of risk that defies simple categorization? To address this, we turned to unsupervised learning approaches to examine the underlying structure of our patient population.

### Unsupervised Learning Uncovers Complex Risk Profiles Rather Than Discrete Patient Clusters

Given the known limitations of EPDS as a screening tool, we conducted comprehensive unsupervised learning analyses to explore the underlying structure of our patient population and identify potential subgroups that may not align with traditional EPDS categorization.

Dimensionality reduction using PCA, t-SNE, and UMAP consistently revealed that patients with elevated postpartum EPDS scores distribute homogeneously throughout the feature space rather than forming distinct clusters. PCA plots are shown in Figure 4 **A**, additional figures can be found in the supplementary material. This pattern persisted across all three ablation dataset configurations (complete, features-dropped, and patients-dropped), indicating that the lack of clear separation is not simply due to the dominance of mental health history features.

This homogeneous distribution suggests several important clinical insights: (1) the relationship between clinical features and postpartum depression risk is highly complex and non-linear, defying simple categorical distinctions; (2) patients at risk for PPD cannot be easily distinguished from low-risk patients using structured clinical variables alone; and (3) multiple subtypes of depression risk profiles likely exist within our cohort, requiring more sophisticated characterization methods beyond the binary and single time-point EPDS categorization.

Optimal cluster determination using silhouette analysis identified 7-9 natural clusters in the data (Figure in the supplementary material) - substantially more than the expected binary categorization and consistent with the multiple error clusters identified in our decision boundary analysis. However, silhouette scores remained low (approximately 0.15), indicating weak cohesion even at optimal cluster counts. Hierarchical clustering dendrograms (Figure 4 **B**) further confirmed the absence of discrete patient subgroups, revealing highly interconnected structures where patients with varying EPDS scores are distributed throughout all branches.

Feature correlation analysis across ablation conditions (Figure 4 **C**) demonstrated that when mental health history is unavailable, demographic and metabolic factors (Black race, government insurance, pregestational diabetes) exhibit the strongest correlations with EPDS scores, though these associations remain modest in magnitude (r < 0.10).

The multiplicity of patient subgroups identified through clustering, combined with the heterogeneous error patterns in our supervised models, supports that the binary outcome for this specific case might be oversimplified. Detailed clustering analyses, including contingency tables, chi-square tests, and cluster purity metrics, are provided in Supplementary Figures.

The absence of discrete patient clusters and the heterogeneous distribution of risk factors presented a significant modeling challenge. Statistical and simple clustering approaches might struggle where the relationship between features and patients cannot be easily stratified into risk categories. This motivated our evaluation of advanced automated machine learning approaches capable of navigating high-dimensional feature spaces and discovering subtle predictive patterns.

### ClinPreAI Agent Matches Specialized Approaches and Outperforms Commercial Solutions Across Data Modalities

We evaluated the ClinPreAI Agent system across five dataset configurations, comparing its performance against traditional AutoML approaches, commercial solutions, large language model baselines, and various traditional models.

In the multimodal configuration (Table 6), the LLM-Enhanced XGBoost approach achieved the highest F1 score of 0.654 *±* 0.013, demonstrating the improvement of effectiveness of combining large language model embeddings with gradient boosting for multimodal clinical data integration relative to other tested approaches. This method leverages Claude Sonnet model to make a pseudo EPDS binary prediction then this is combined with structured features in an XGBoost classifier. While this specialized approach excelled at multimodal integration, we also evaluated whether a more generalizable agentic system could achieve competitive performance without task-specific architectural design.

The ClinPreAI Agent achieved competitive performance against other methods on this configuration with F1 = 0.648 *±* 0.043, substantially outperforming commercial AutoML solutions and traditional baselines while demonstrating greater flexibility across diverse data modalities. The agent’s autonomous approach to feature engineering and model selection proved particularly effective for integrating different data sources, though with somewhat higher variance compared to the more specialized LLM-XGBoost pipeline.

In comparison, the non-agentic ClinPreAI AutoML framework achieved F1 = 0.585 *±* 0.014 on the same multimodal data—a fair score that fell short of both the agent and LLM-XGBoost approaches, as well as its own structured-only performance (F1 = 0.640 *±* 0.019, Table 4). This performance gap suggests that the AutoML pipeline could benefit from enhanced preprocessing strategies for clinical narratives, such as improved text chunking, summarization or improved embedding approaches. Notably, AWS Canvas (both Standard and Quick modes) failed to process the multimodal dataset entirely, likely due to computational limitations when handling extensive clinical text, underscoring scalability challenges in commercial AutoML platforms for complex healthcare applications.

**Table 4:** Performance Metrics on Structured Dataset.

| Method | F1 Score | Accuracy | Precision | Recall | ROC-AUC |
| --- | --- | --- | --- | --- | --- |
| Random Forest | 0.408 ± 0.000 | 0.690 ± 0.000 | 0.345 ± 0.000 | 0.500 ± 0.000 | 0.685 ± 0.023 |
| Stratified Ensemble | 0.439 ± 0.029 | 0.679 ± 0.018 | 0.481 ± 0.031 | 0.409 ± 0.051 | 0.651 ± 0.017 |
| ClinPreAI AutoML | 0.640 ± 0.019 | 0.696 ± 0.015 | 0.642 ± 0.019 | 0.638 ± 0.019 | 0.657 ± 0.021 |
| AWS Canvas Quick | 0.538 ± 0.000 | 0.681 ± 0.000 | 0.489 ± 0.000 | 0.599 ± 0.000 | 0.668 ± 0.000 |
| AWS Canvas Standard | 0.550 ± 0.000 | 0.698 ± 0.000 | 0.512 ± 0.000 | 0.595 ± 0.000 | 0.690 ± 0.000 |
| ClinPreAI Agent | **0.678 ± 0.028** | 0.670 ± 0.032 | 0.802 ± 0.008 | 0.690 ± 0.069 | 0.688 ± 0.007 |

We next examined whether strong performance could be maintained when restricting models to structured clinical variables without clinical narratives (Table 4), the ClinPreAI Agent achieved the highest performance with F1 = 0.678 *±* 0.028, demonstrating robust handling of traditional tabular healthcare data. The ClinPreAI AutoML approach performed competitively on this configuration with F1 = 0.640 *±* 0.019, though with slightly lower mean performance.

Commercial and traditional baselines showed substantially weaker performance. AWS Canvas Standard configuration reached F1 = 0.550, while the Quick configuration achieved F1 = 0.538. Most concerning, the default Random Forest baseline severely underperformed with F1 = 0.408, failing to exceed even the naïve majority-class predictor. This failure across multiple traditional approaches indicates fundamental limitations in handling class imbalance without careful algorithm selection and hyperparameter tuning—precisely the problem that automated approaches like ClinPreAI are designed to address.

To assess the independent predictive value of clinical narratives, we evaluated models using only social worker notes or extracted HPO terms (Table 5); these text-only models performed worse than structured-only or multimodal approaches. The best text-only result came from the ClinPreAI Agent (F1 = 0.628 *±* 0.012), though with substantial variance across runs. Claude 0-shot language models provided more stable results, with Claude Sonnet achieving F1 = 0.566 *±* 0.003 and Claude Opus reaching F1 = 0.568 *±* 0.005. The ClinPreAI AutoML approach yielded F1 = 0.533 *±* 0.028, while traditional text embeddings with logistic regression produced the weakest performance (F1 = 0.447 *±* 0.014).

**Table 5:** Performance Metrics on Text Dataset.

| Method | F1 Score | Accuracy | Precision | Recall | ROC-AUC |
| --- | --- | --- | --- | --- | --- |
| ClinPreAI AutoML | 0.533 ± 0.028 | 0.573 ± 0.030 | 0.536 ± 0.027 | 0.536 ± 0.029 | 0.564 ± 0.042 |
| Text Embeddings | 0.447 ± 0.014 | 0.651 ± 0.009 | 0.594 ± 0.054 | 0.515 ± 0.008 | 0.603 ± 0.024 |
| ClinPreAI Agent | **0.628 ± 0.012** | 0.620 ± 0.011 | 0.638 ± 0.012 | 0.620 ± 0.011 | — |
| Claude Opus | 0.568 ± 0.005 | 0.693 ± 0.004 | 0.554 ± 0.006 | 0.583 ± 0.004 | 0.667 ± 0.004 |
| Claude Sonnet | 0.566 ± 0.003 | 0.657 ± 0.004 | 0.504 ± 0.004 | 0.645 ± 0.006 | 0.654 ± 0.003 |
| Claude Sonnet Reasoning | 0.545 ± 0.030 | 0.680 ± 0.013 | 0.533 ± 0.019 | 0.557 ± 0.042 | 0.650 ± 0.019 |

**Table 6:** Performance Metrics on Multimodal Dataset.

| Method | F1 Score | Accuracy | Precision | Recall | ROC-AUC |
| --- | --- | --- | --- | --- | --- |
| Heuristic | 0.502 ± 0.011 | 0.698 ± 0.008 | 0.514 ± 0.013 | 0.492 ± 0.013 | 0.641 ± 0.008 |
| ClinPreAI AutoML | 0.585 ± 0.014 | 0.704 ± 0.017 | 0.641 ± 0.032 | 0.585 ± 0.013 | 0.674 ± 0.023 |
| ClinPreAI Agent | 0.648 ± 0.043 | 0.638 ± 0.048 | 0.700 ± 0.013 | 0.638 ± 0.048 | 0.688 ± 0.009 |
| LLM-XGBoost | **0.654 ± 0.013** | 0.691 ± 0.009 | 0.673 ± 0.012 | 0.691 ± 0.009 | 0.681 ± 0.029 |
| Claude Opus | 0.511 ± 0.001 | 0.599 ± 0.001 | 0.423 ± 0.001 | 0.645 ± 0.002 | 0.611 ± 0.001 |
| Claude Sonnet | 0.510 ± 0.001 | 0.586 ± 0.002 | 0.415 ± 0.001 | 0.664 ± 0.002 | 0.606 ± 0.001 |
| Claude Sonnet Reasoning | 0.517 ± 0.015 | 0.607 ± 0.014 | 0.420 ± 0.018 | 0.672 ± 0.014 | 0.625 ± 0.013 |
| Qwen Original | 0.208 ± 0.018 | 0.666 ± 0.005 | 0.341 ± 0.017 | 0.150 ± 0.016 | — |
| Qwen Finetuned | 0.069 ± 0.070 | 0.706 ± 0.006 | 0.485 ± 0.328 | 0.041 ± 0.045 | — |

It is important to note that the majority class baseline for the text only yields F1 = 0.529 due to dropping patients without available social worker notes. Critically, all text-only models barely outperform the majority-class predictor, suggesting that extracting actionable predictive signal from clinical narratives requires more sophisticated processing than zero-shot inference or simple embedding approaches.

We evaluated two configurations incorporating engineered latent features: Latent Multimodal v1 (adding dimensionality reduction, unsupervised clustering, and statistical interactions) and Latent Multimodal v2 (further augmented with domain-specific HPO-derived risk scores).

For Latent Multimodal v1 (Table 7), ClinPreAI AutoML achieved F1 = 0.627 *±* 0.010, showing improvement over baseline multimodal results. However, the ClinPreAI Agent’s performance declined to F1 = 0.624 *±* 0.048 with substantially increased variance. Qualitative analysis of the agent’s execution logs revealed that it generated diverse planning strategies for incorporating latent features across different runs, resulting in inconsistent approaches. In several cases, the agent’s plans became overly complex, leading to execution failures—a limitation of the current architecture that highlights opportunities for improved planning module constraints.

**Table 7:** Performance Metrics on Latent Multimodal v1 Dataset.

| Method | F1 Score | Accuracy | Precision | Recall | ROC-AUC |
| --- | --- | --- | --- | --- | --- |
| ClinPreAI AutoML | **0.627 ± 0.010** | 0.701 ± 0.015 | 0.643 ± 0.017 | 0.622 ± 0.008 | 0.679 ± 0.020 |
| ClinPreAI Agent | 0.624 ± 0.048 | 0.620 ± 0.053 | 0.666 ± 0.016 | 0.620 ± 0.053 | 0.641 ± 0.010 |

For Latent Multimodal v2 (Table 8), which added HPO-derived engineered features, the ClinPreAI Agent recovered to F1 = 0.652 *±* 0.031, while ClinPreAI AutoML maintained stable performance at F1 = 0.621 *±* 0.011. While the agent’s score appeared slightly higher, the overlapping confidence intervals suggest this difference may reflect variance rather than true superiority. These results indicate that domain-specific feature engineering can enhance model performance, though the optimal integration strategy remains architecture-dependent.

**Table 8:** Performance Metrics on Latent Multimodal v2 Dataset.

| Method | F1 Score | Accuracy | Precision | Recall | ROC-AUC |
| --- | --- | --- | --- | --- | --- |
| ClinPreAI AutoML | 0.621 ± 0.011 | 0.702 ± 0.016 | 0.643 ± 0.017 | 0.616 ± 0.010 | 0.675 ± 0.014 |
| ClinPreAI Agent | **0.652 ± 0.031** | 0.644 ± 0.034 | 0.684 ± 0.008 | 0.644 ± 0.034 | 0.661 ± 0.017 |

To contextualize our results within the broader landscape of AI approaches, we evaluated several state-of-the-art large language models, including Claude 4 Opus, Claude 4 Sonnet (with and without extended reasoning), and Qwen2.5-7B (zero-shot and fine-tuned). These models were assessed on both text-only (Table 5) and multimodal configurations (Table 6) across five independent runs.

Zero-shot large language models demonstrated limited effectiveness for this clinical prediction task. On text-only data, Claude variants achieved F1 scores in the range of 0.524-0.568, inferior to the ClinPreAI Agent approach. Counterintuitively, incorporating structured clinical data alongside text led to degraded rather than improved performance across all Claude models (multimodal F1: 0.510-0.517), suggesting that table serialization for zero-shot inference introduced noise or formatting challenges that hindered effective multimodal integration.

The smaller Qwen2.5-7B model performed markedly worse, achieving F1 = 0.208 *±* 0.018 in zero-shot mode and only F1 = 0.069 *±* 0.018 after fine-tuning with QLoRA—both substantially below baseline. The high variance in fine-tuned performance (standard deviation = 0.070) indicates unstable learning, likely due to limited model capacity and lack of domain-specific pretraining on clinical vocabularies.

These findings underscore that general-purpose language models, even frontier systems like Claude 4, require substantial adaptation and optimized prompt engineering for specialized clinical prediction tasks. This performance gap may reflect two non-mutually exclusive factors. First, clinical text data contains substantial noise that may limit its direct predictive utility for specific outcomes. Second, our zero-shot implementation may not adequately handle large-scale clinical text; more sophisticated approaches incorporating chunking, summarization, or hierarchical information extraction may be necessary to distill clinically relevant signals from verbose documentation.

Figure 2 presents comprehensive comparisons across all modeling strategies and dataset configurations. The top performing systems were: (1) LLM-Enhanced XGBoost on Multimodal data (F1 = 0.654 *±* 0.013), (2) ClinPreAI Agent on Structured data (F1 = 0.678 *±* 0.028), and (3) ClinPreAI Agent on Multimodal data (F1 = 0.648 *±* 0.043).

While LLM-XGBoost achieved the highest score on multimodal integration, the ClinPreAI Agent demonstrated consistent strong performance across all configurations, including matching the top F1 score on structured data and achieving competitive results on latent feature configurations. This consistent performance across diverse data modalities demonstrates that agentic approaches excel at both multimodal integration and structured data analysis, with the flexibility to adapt to various input types without task-specific architectural modifications.

The specialized LLM-XGBoost approach highlights the value of tailored strategies for multimodal clinical data, though its evaluation was limited to the multimodal configuration. Traditional AutoML frameworks remained competitive for well-structured tabular data, while commercial solutions, general-purpose large language model baselines, and traditional approaches showed substantially weaker performance. The consistent superiority of ClinPreAI methods over these baselines validates the value of task-agnostic automated machine learning for clinical prediction, with both agentic and specialized approaches offering complementary strengths.

While these results demonstrated the agentic AI system strong relative performance against other methods, we wanted to learn what was the critical features the models were considering to make the classification decision and if they were different than the statistical differences between the groups found in the cohort characterization. Understanding this dependency is essential for determining whether our approach identifies truly novel risk factors or primarily flags patients with anxiety or depression history.

### Mental Health History Dominates Predictive Performance

The ablation study revealed performance degradation corresponding to progressive removal of mental health features (Figure 3). The optimized XGBoost model achieved substantial discriminative capability on the complete dataset (F1=0.68), comparable to performance obtained through the Agent approach for model selection. The relative performance gains across ablation conditions are visualized in Figure 3 **B**. On the complete dataset, the optimized XGBoost model achieved 20.2% improvement over the majority class baseline. This gain decreased to 9.6% when special features were removed, and further diminished to only 1.9% when all mental health features were excluded, demonstrating the critical importance of these variables for predictive performance.

When the two key depression-related features (prior anxiety/depression diagnosis and pharmacological treatment) were removed, model performance declined to F1 = 0.56. This substantial but not catastrophic degradation indicates that while mental health history dominates prediction, alternative clinical markers retain modest prognostic value. Feature importance analysis (Figure 3 **C**) revealed that the model compensated by leveraging proximal indicators: chronic medical conditions emerged as the leading feature in this configuration, followed by sociodemographic factors including insurance type and marital status.

In contrast, excluding patients with documented mental health history entirely (patients-dropped configuration) collapsed model performance to near-random levels, with F1 scores comparable to stratified baseline prediction across all models tested (other models like logistic regression and random forest metrics are shown in the supplementary material). This decline exposes a critical limitation: the predictive signal in our dataset derives overwhelmingly from recurrence risk among patients with established mental health vulnerabilities.

Supplementary analyses show degradation across all metrics (accuracy, precision, recall, F1); in the patients-dropped setting, even a majority-class baseline matches optimized models, indicating no learnable signal. This reveals that our current structured clinical data struggles to identify de novo postpartum depression risk without prior mental-health documentation—models largely capture recurrence and add little value for primary-prevention screening; full metrics and confusion matrices are provided in the supplementary material.

To further investigate whether de novo risk factors for postpartum depression could be identified independent of established mental health history, we conducted a complementary prediction task targeting prior anxiety/depression diagnosis as the outcome variable (see Supplementary Material). This reverse prediction task revealed patterns consistent with our primary findings: when the full feature set including EPDS label and pharmacological treatment history was available, prior anxiety diagnosis was highly predictable (F1 > 0.74). However, systematic removal of depression-related features resulted in severe performance degradation, with F1 scores declining to 0.63 and approaching random classification levels. This mirrors the performance trajectory observed in our primary ablation studies and confirms the strong interdependency among mental health indicators in our dataset. The symmetric degradation patterns across both prediction directions (EPDS from features vs. anxiety history from features) reinforce our conclusion that current clinical data structures capture primarily recurrence-related signals rather than de novo risk pathways, as discussed in subsequent sections addressing feature correlation.

The ablation study revealed that our models largely captured recurrence risk among patients with prior mental health diagnoses. The lack of new identifiable risk factors for PPD and the absolute low performance of the best performing model prompted us to examine error analysis systematically. To understand if it was a limitation of our modeling approaches or our study design.

### Automated Error Analysis Identifies Decision Boundaries and Demographic Disparities

The ClinPreAI AutoML framework extends beyond model selection to provide comprehensive interpretability through systematic error analysis. This integrated approach enables researchers to not only achieve optimal predictive performance but also understand model behavior, identify potential biases, and develop targeted refinement strategies. Some of these plots are shown in Figure 5.

#### Decision Boundary Characterization

Analysis of prediction confidence revealed a clear relationship between model uncertainty and error occurrence (Figure 5 **A**,**B**). Samples near the classification decision boundary (<10th percentile distance) exhibited substantially elevated error rates of approximately 60%, compared to ∼ 30% for high-confidence predictions. PCA projection of the feature space (Figure 5 **C**) revealed distinct distributional patterns between correct and erroneous predictions. Error samples (red) exhibited higher density in the upper left quadrant with limited variance along the PC1 axis, whereas correctly classified samples (green) demonstrated greater dispersion along PC1, suggesting that errors cluster in a restricted region of the feature space while correct predictions span a broader representational range.

#### Confidence Calibration

Model confidence analysis (Figure 5 **D**,**E**) demonstrated acceptable calibration, with predicted probabilities closely tracking actual positive class frequencies along the ideal diagonal. High-confidence predictions (≥0.8) maintained substantially lower error rates compared to low-confidence predictions (<0.6), validating the utility of confidence scores for risk-stratified clinical workflows. The analysis revealed an asymmetry in error types, with Type II errors (false negatives, missed cases) occurring more frequently than Type I errors (false positives, over-diagnosis) shown in Figure 5 **F**, suggesting potential opportunities for threshold optimization favoring sensitivity over specificity in clinical deployment.

#### Feature-Specific Error Patterns

Examination of feature-error correlations (Figure 5 **G**) identified systematic relationships between patient characteristics and prediction failures. Race emerged as significantly correlated with error patterns, with Black race showing positive correlation with Type I errors and Hispanic/Latino ethnicity demonstrating negative correlation. Gestational age at admission exhibited negative correlation with Type II errors, indicating improved model performance for pregnancies with longer durations at the time of hospitalization. These patterns highlight potential demographic disparities requiring targeted data collection strategies for model refinement.

The combined error analysis framework provides several advantages for clinical machine learning applications:

1. **Targeted Clinical Review**: Samples near decision boundaries can be flagged for additional clinical assessment, optimizing resource allocation for cases where algorithmic predictions are most uncertain.
2. **Bias Detection and Mitigation**: Feature-specific error analysis enables proactive identification of demographic disparities, supporting equity-focused model refinement and deployment strategies.
3. **Confidence-Based Care**: Calibrated confidence scores support risk-stratified workflows, allowing clinicians to prioritize high-risk cases while applying less intensive monitoring to low-risk predictions.
4. **Iterative Improvement Guidance**: Detailed error clustering identifies specific patient subpopulations requiring additional data collection or alternative modeling approaches.

These automated error analyses provided valuable statistical insights into model behavior and potential biases. However, statistical patterns alone cannot fully capture the clinical validity of predictions or reveal whether errors reflect true model failures versus the inherent difficulty of prospective prediction. To address this gap, we complemented our quantitative analyses with detailed qualitative review by clinical experts.

### Clinical Expert Review Validates Model Reasoning Despite Prediction Errors

To complement automated statistical analysis, we conducted two independent qualitative assessments of model predictions: (1) a detailed case-by-case review of misclassified cases (n=15) with systematic Type I and Type II error pattern analysis, and (2) an independent expert clinical validation by a perinatal mental health specialist reviewing a separate sample (n=20). These qualitative examinations revealed important insights into prediction errors that extend beyond patterns identifiable through statistical methods alone and provided external validation of the model’s clinical reasoning.

#### Detailed Error Pattern Analysis (n=15)

We performed systematic manual review of 15 representative misclassified cases (9 Type I errors and 6 Type II errors) to understand the nature and underlying causes of prediction failures. Model-generated summaries appeared clinically reasonable and consistent with conclusions that would be drawn from direct review of the medical records. *Type I Error Patterns (False Positives)*. Manual inspection of false positive cases revealed a consistent pattern wherein model predictions aligned with documented clinical risk factors, yet ground-truth EPDS scores remained low. In 8 of 9 Type I error cases, the LLM-based feature extraction and subsequent model predictions appeared clinically justified based on available documentation. For instance, Case 0489 presented with extensive social worker notes documenting feelings of guilt, sadness, and depression (“not being a good mom by being away from her children”), psychological support utilization, and explicit documentation of depressed affect. Despite this substantial evidence suggesting elevated postpartum depression risk, the patient’s postpartum EPDS score was below threshold. Similarly, multiple false positive cases involved patients with documented histories of anxiety and depression, pharmacologic treatment, relevant HPO terms (anxiety, depressivity, cognitive deficits), and psychosocial complexity factors including unemployment, transportation dependency, and child proactive services (CPS)/legal involvement. *Type II Error Patterns (False Negatives)*. Type II errors predominantly occurred in patients presenting with apparently lower-risk profiles based on prenatal documentation. Three of six false negative cases exhibited sparse clinical documentation, with several lacking available social worker notes entirely. These patients frequently demonstrated demographic characteristics associated with lower risk (predominantly White race, married status, private insurance) despite documented chronic medical conditions. Notably, in Case 0490, financial stress and anxiety were documented but the model assigned medium confidence to its negative prediction, suggesting insufficient weighting of psychosocial stressors in the feature representation.

#### Independent Expert Clinical Validation (n=20)

To provide external validation and reduce potential reviewer bias, a perinatal mental health specialist independently evaluated model predictions on a separate sample of 20 cases without knowledge of our initial error analysis. The expert reviewer had access to the full medical records, model predictions, and ground-truth EPDS outcomes. The expert agreed with model predictions in 15 of 20 cases (75% concordance). Importantly, even in cases of disagreement, the expert noted that the model’s reasoning appeared clinically valid and that other clinicians would likely find the predictions reasonable. The expert emphasized the inherent difficulty of prospectively predicting postpartum depression, acknowledging that significant intervening events (e.g., additional pregnancy complications, delivery, immediate postpartum experiences) occur between data collection and outcome assessment, making this “a very hard machine learning problem” even for experienced clinicians. The expert was particularly impressed with the quality and clinical relevance of the LLM-generated summaries, noting they were “reasonable and aligned with what I myself would have written after review of the notes.” This observation supports potential applications of LLM-based summarization tools for clinical workflows, where efficient extraction of relevant information from extensive documentation remains a significant time burden for healthcare providers.

#### Synthesis and Clinical Implications

Both independent reviews converged on several key findings. First, model predictions demonstrated clinical validity and reflected reasoning patterns consistent with expert clinical judgment. Second, many prediction errors appeared to result from the fundamental challenge of prospective prediction rather than systematic model failures—patients with documented risk factors may not develop postpartum depression, while others with apparently lower-risk profiles may experience elevated symptoms due to factors not captured in prenatal documentation. Third, the quality of LLM-generated clinical summaries suggests broader potential for AI-assisted medical record review beyond prediction tasks. These findings underscore both the clinical interpretability of our approach and the inherent complexity of predicting postpartum mental health outcomes from prenatal data alone.

## Discussion

This study demonstrates that autonomous agentic AI systems can effectively develop clinical prediction models that match or exceed specialized human-designed approaches. ClinPreAI achieved F1 scores of 0.65-0.68 across diverse data modalities, substantially outperforming traditional AutoML (F1: 0.59-0.64), commercial platforms (AWS Canvas F1: 0.54-0.55), and zero-shot large language models (Claude Opus F1: 0.51-0.52). However, our findings also reveal a critical limitation: current approaches for PPD prediction rely overwhelmingly on documented mental health history, offering limited utility for primary prevention in patients without prior psychiatric documentation. This limitation has also been noted by prior studies in the field[47].

The application of LLM based approaches—both the specialized LLM-XGBoost method (F1: 0.654 *±* 0.013 on multimodal data) and the ClinPreAI Agent (F1: 0.678 *±* 0.028 on structured data)—stems from their ability to effectively integrate heterogeneous data types and adapt to dataset characteristics. The LLM-XGBoost method’s success demonstrates the value of specialized strategies for combining structured and unstructured clinical data, particularly leveraging LLM predictions as intermediate representations. Meanwhile, the ClinPreAI Agent’s consistent strong performance across all configurations highlights advantages of autonomous planning: systematic exploration of feature engineering strategies tailored to specific datasets, self-verification through debugging modules enabling error correction without human intervention, and iterative refinement through feedback integration.

Compared to prior PPD prediction models [24, 25, 47], our approach uniquely operates on single-encounter data from first antepartum hospitalization rather than delivery-time or longitudinal prenatal data. While this temporal constraint increases prediction difficulty, it enables earlier risk stratification with greater clinical benefit. Our F1 scores (0.65-0.68) compare favorably to baseline models, particularly given the challenging single-timepoint prediction task and class imbalance in our cohort.

However, ablation studies exposed a fundamental limitation in the structured data signal. When patients with documented mental health history were excluded, model performance collapsed to near-baseline levels across all algorithms tested—including optimized XGBoost, logistic regression, and random forests. This uniform degradation indicates the limitation stems from available feature space rather than modeling approach. Current structured EHR data capture primarily recurrence-related signals (patients with prior depression/anxiety developing PPD) rather than de novo risk pathways. Unsupervised clustering analyses reinforced this finding, revealing continuous rather than discrete risk stratification and suggesting that binary EPDS categorization may oversimplify depression phenotype heterogeneity.

For clinical implementation, our findings suggest several considerations. First, agentic AI systems like ClinPreAI can effectively democratize access to sophisticated predictive modeling, enabling domain experts without ML training to develop robust clinical tools. Also, the ClinPreAI AutoML framework’s comprehensive interpretability outputs (SHAP values, error analysis, feature importance) provide transparency essential for healthcare data analytical applications. Second, institutions implementing PPD screening should recognize that prediction models trained on structured EHR data will perform best for patients with documented mental health history—a population already identified as high-risk through simple heuristics.

The high concordance between model predictions and independent expert clinical assessment—despite classification errors relative to ground-truth EPDS scores—suggests potential applications beyond binary prediction. LLM-generated clinical summaries demonstrated high quality and clinical relevance, suggesting utility for extracting and synthesizing information from extensive documentation even when prediction accuracy is limited. This capability could reduce clinician time burden for medical record review and support clinical decision-making through information synthesis rather than deterministic classification.

### Study Design and Measurement

This single-center retrospective study limits generalizability, though our diverse patient population enhances external validity. The substantial class imbalance (31% positive cases) and moderate overall performance indicate that even our best models misclassify approximately one-third of cases, raising questions about clinical utility thresholds for implementation.

A key limitation of this work may lie less in model performance than in the quality, timing, and scope of the outcome labels— particularly because postpartum EPDS was measured at a single timepoint. This design choice limits our ability to capture and predict delayed symptom onset, so some apparent “misses” may reflect cases that emerge after the measurement window rather than true negatives at the time of assessment. In this context, the high concordance between model predictions and independent expert clinical assessment—despite classification errors relative to EPDS thresholds—suggests that some apparent failures may partly reflect limitations in the ground-truth labels.

Three mechanisms may contribute to discordance between clinical documentation and self-reported EPDS scores. First, *temporal misalignment*: intervening events between prenatal data collection and postpartum outcome measurement (e.g., delivery complications, neonatal outcomes, or early postpartum stressors) can substantially shift symptom trajectories, yet these exposures are not represented in the prenatal feature space and therefore function as unobserved confounders. Second, *treatment-effect confounding*: documentation of risk factors or emerging symptoms may trigger clinical actions (e.g., counseling, psychotherapy referral, pharmacotherapy initiation or adjustment, increased follow-up, or social support interventions) that reduce symptom severity before EPDS screening. As a result, individuals correctly flagged as high risk may score below the EPDS threshold at the postpartum assessment, creating apparent “false positives” that reflect successful symptom control rather than erroneous prediction. Conversely, those without documented risk may receive less monitoring and fewer opportunities for early intervention, which can exacerbate apparent discordance in the opposite direction. Because treatment initiation, adherence, dose changes, and care engagement were not available, we cannot quantify the magnitude of this effect or determine whether discordant cases represent misclassification versus clinical response to earlier risk identification. Third, *self-report limitations* inherent to EPDS measurement—including stigma, social desirability bias, and underreporting—may lead to systematically lower scores that diverge from clinician observations.

Together, these considerations suggest that disagreement between the model and EPDS may reflect measurement limitations rather than model error, though separating these sources remains difficult without gold-standard psychiatric diagnostic assessments and longitudinal follow-up. Clinically, accurately identifying the *initial* prenatal depression episode also remains challenging and was not addressed in this study; developing reliable methods to detect this event is an important future direction, given its potential value for predicting later postpartum symptom trajectories.

### Technical Considerations

The ClinPreAI Agent exhibited high variance in planning strategies and occasional execution failures when incorporating complex engineered features, particularly in the Latent Multimodal configurations. These technical limitations highlight architectural constraints requiring refinement before clinical deployment. The system’s autonomous nature, while enabling exploration of novel approaches, introduces high variable approaches that may concern clinicians accustomed to transparent, deterministic tools. Careful consideration of human validation, fail-safe protocols, and model reproducibility would be essential for clinical implementation.

Several complementary strategies could address current limitations. *Data enrichment approaches* should explore additional information sources beyond traditional EHR variables, including patient-reported outcome measures collected during antepartum hospitalization, structured assessments of social support and psychosocial stressors, and longitudinal data incorporating delivery and immediate postpartum information. *Validation studies* in diverse healthcare settings are essential to establish generalizability and assess real-world clinical utility, particularly in populations with different demographic compositions and practice patterns.

*Architectural improvements* to the ClinPreAI Agent could address current variance and execution challenges. Integration of retrieval-augmented generation (RAG) would enable the system to maintain memory of successful approaches across iterations, reducing redundant experimentation. Adoption of the Model Context Protocol (MCP) would expand the agent’s toolbox, enabling access to specialized medical ontologies and domain-specific computational resources. Prompt optimization strategies targeting the planning module could increase both performance consistency and solution quality. LLM latent feature mining and self improvement, could also enhance model performance. These enhancements could reduce high variance while maintaining the flexibility during code execution.

Finally, *methodological advances* should address outcome measurement limitations through prospective studies incorporating structured psychiatric diagnostic interviews as gold-standard outcomes, enabling assessment of whether model-label discordance reflects prediction limitations versus measurement error. Development of continuous rather than binary outcome measures could better capture depression phenotype heterogeneity revealed by unsupervised analyses. At the same time, we formulated prediction as a binary task for interpretability and operational utility (e.g., risk stratification and threshold-based decision support) and because binary targets are straightforward to train and evaluate; while extending to continuous EPDS prediction is feasible, our results suggest that signal is already modest even under dichotomization, and predicting the full continuous score may not improve—and could even reduce—predictive performance due to added measurement noise and within-class heterogeneity.

This work represents a paradigm shift in PPD prediction from static AutoML frameworks toward adaptive autonomous systems capable of iterative refinement and self-correction. The ClinPreAI Agent’s modular architecture—comprising research, planning, coding, debugging, and interpretation modules—provides a generalizable template for deploying AI agents across diverse clinical prediction tasks. By automating the iterative process of experimentation, debugging, and refinement, agentic systems democratize access to sophisticated predictive modeling. This is particularly valuable in clinical settings where domain experts may lack ML training, and in complex multimodal scenarios where optimal modeling strategies are non-obvious.

The enhanced interpretability provided through comprehensive error analysis, detailed manual case inspection, and explicit reasoning traces demonstrates that AI systems can maintain transparency essential for healthcare applications. The high concordance between model predictions and independent expert clinical assessment validates the clinical reasoning captured by these systems, suggesting broader applications beyond binary classification—including clinical summarization, information synthesis, and decision support.

To the best of our knowledge, this represents the first application of agentic AI systems to perinatal mental health prediction, establishing a foundation for broader deployment of autonomous AI agents in clinical medicine. While challenges remain—including architectural refinements to reduce variance, validation in diverse populations, and improved outcome measurement—agentic AI systems offer compelling proof-of-concept that sophisticated clinical prediction tools can be developed and deployed by domain experts without specialized machine learning expertise, potentially transforming how predictive models are created and implemented in healthcare settings.

## Method

### Study Design and Population

This retrospective cohort study included pregnant individuals hospitalized for management of medical or obstetric complications at a large referral center: Texas Children’s Hospital Pavilion for Women serving a diverse population from March 2012 to March 2025. Data were extracted from electronic health records, including demographics, medical history, social worker notes, and postpartum Edinburgh Postnatal Depression Scale (EPDS) scores collected within 8 weeks of delivery.

The primary outcome was a postpartum EPDS score ≥ 10, indicating clinically significant PPD symptoms. From an initial cohort of 6,643 patients, we excluded those without available EPDS scores, resulting in a final analytical sample of 4,161 patients. Among these, 1,291 (31.0%) had EPDS scores ≥ 10, while 2,870 (69.0%) had scores *<* 10.

### Data Sources and Processing

We extracted 27 structured variables from the EHR, categorized across five clinical domains: demographics (maternal age, race, Hispanic/Latino ethnicity, Spanish language preference, marital status, insurance type), obstetric history (gestational age on admission, hospitalization length, multiparous status), psychiatric history (prior anxiety or depression diagnosis, and prior pharmacologic treatment for anxiety or depression), medical history (pregestational diabetes, chronic hypertension, multiple chronic medical conditions), and pregnancy complications (preterm premature rupture of membranes, placenta accreta spectrum, preeclampsia, gestational diabetes, multiple gestation). Table 3 provides detailed variable descriptions. We summarized cohort characteristics using means (SD) or medians (IQR) for continuous variables and counts (percentages) for categorical variables, assessed distributional assumptions (Shapiro–Wilk/Kolmogorov–Smirnov; Levene’s), and compared EPDS groups (*<*10 vs. ≥ 10) using t-tests or Mann– Whitney U tests for continuous variables and *χ*^2^ or Fisher’s exact tests for categorical variables, as appropriate.

Structured data underwent systematic transformation. Medication records were classified to identify antidepressant or antianxiety treatments prior to first hospital admission. Postpartum EPDS scores were processed to extract the highest recorded value per patient, with scores ≥ 10 classified as positive for depression risk. Categorical variables were grouped into clinically relevant categories, e.g., different insurance providers were classified into Private, Government, and Self-pay categories. Similar transformations were applied for race and gestational age, which were then converted into binary indicators. All data sources were aggregated by medical record number to create unified patient-level datasets. To avoid duplication, only the first hospital admission encounter was retained for each patient; subsequent admissions or pregnancies were not included in the analysis.

We included social worker notes authored by social workers and licensed clinical social workers, as these documents contained valuable information about socioeconomic factors and mental health discussions. This selective filtering reduced noise and provided higher quality textual data for analysis. Notes from multiple time periods during the first admission were concatenated for each patient, and text was preprocessed using standard NLP techniques including tokenization, lemmatization, and stop-word removal while preserving domain-specific medical terminology using the NLTK python library.

Human Phenotype Ontology (HPO) annotation was applied to extract standardized phenotypic terms and codes using FastHPOCR [49].

### Feature Engineering and Latent Feature Mining

The feature engineering pipeline expanded our feature space from 27 to over 200 variables through multiple complementary approaches:

#### Dimensionality Reduction and Latent Factors

We applied Principal Component Analysis (extracting 10 components preserving majority variance), Factor Analysis (identifying 5 underlying latent constructs), Independent Component Analysis (decomposing features into 5 statistically independent components), and Non-negative Matrix Factorization (extracting 5 additive components representing cumulative risk factors).

#### Unsupervised Pattern Discovery

We implemented K-means clustering at multiple granularities (k=3, 5, 7), retaining both cluster assignments and distances to centroids; DBSCAN to identify outlier patients and rare clinical presentations; and t-SNE embeddings creating 2-dimensional representations preserving local neighborhood structures.

#### Statistical and Interaction Features

We systematically generated interaction terms between the top 10 highest-variance features; ratio and difference features for feature pairs; z-scores, percentile ranks, and median distances for each numeric feature; and row-wise statistical summaries (mean, standard deviation, skewness, kurtosis, range, IQR).

#### Domain-Specific Risk Aggregation

Leveraging clinical domain knowledge, we developed a comprehensive framework for extracting and aggregating mental health risk indicators from social worker notes using HPO terms. We implemented pattern-based extraction focusing on six primary symptom domains: depression (22 HPO terms), anxiety (18 terms), behavioral abnormalities (16 terms), sleep/fatigue disturbances, cognitive symptoms, and pregnancy-specific indicators. The extraction framework employed context-aware processing including negation detection, proximity-based symptom association, and pregnancy-specific terminology adaptation.

We engineered multiple risk aggregation strategies: (1) Mental Health Severity Score—a composite metric calculated as the sum of depression, anxiety, and behavioral term counts (range: 0 to unlimited, with scores ≥ 3 indicating high symptom burden); (2) Binary Presence Features for each symptom domain; (3) Count-Based Features quantifying distinct terms per category; and (4) Risk Stratification Indicators including suicidal risk assessment and depression-anxiety comorbidity flags. Evidence-based clinical decision rules incorporated multiple aggregation strategies (sum, mean, maximum, standard deviation) across features. Complete methodology, including detailed extraction patterns, the full HPO term dictionary with 90+ terms, clinical reasoning logic, and validation procedures, is documented in Supplementary Material.

#### Dataset Configurations

We created five dataset configurations for comparative analysis: (1) **Structured Only**—original 27 structured clinical variables; (2) **Text Only**—social worker notes and extracted HPO terms; (3) **Multimodal**—combined structured and text data; (4) **Latent Multimodal v1**—multimodal data enhanced with latent features from dimensionality reduction, unsupervised pattern discovery, and statistical interactions; (5) **Latent Multimodal v2**—Latent Multimodal v1 plus domain-specific risk aggregation features.

### ClinPreAI Agent Architecture

We developed an agentic AI system that autonomously designs, implements, and evaluates machine learning solutions for clinical prediction tasks. The system architecture (Figure 1) consists of five specialized modules working in an iterative pipeline to progressively refine predictive models through autonomous experimentation and user feedback integration.

**Figure 1.**
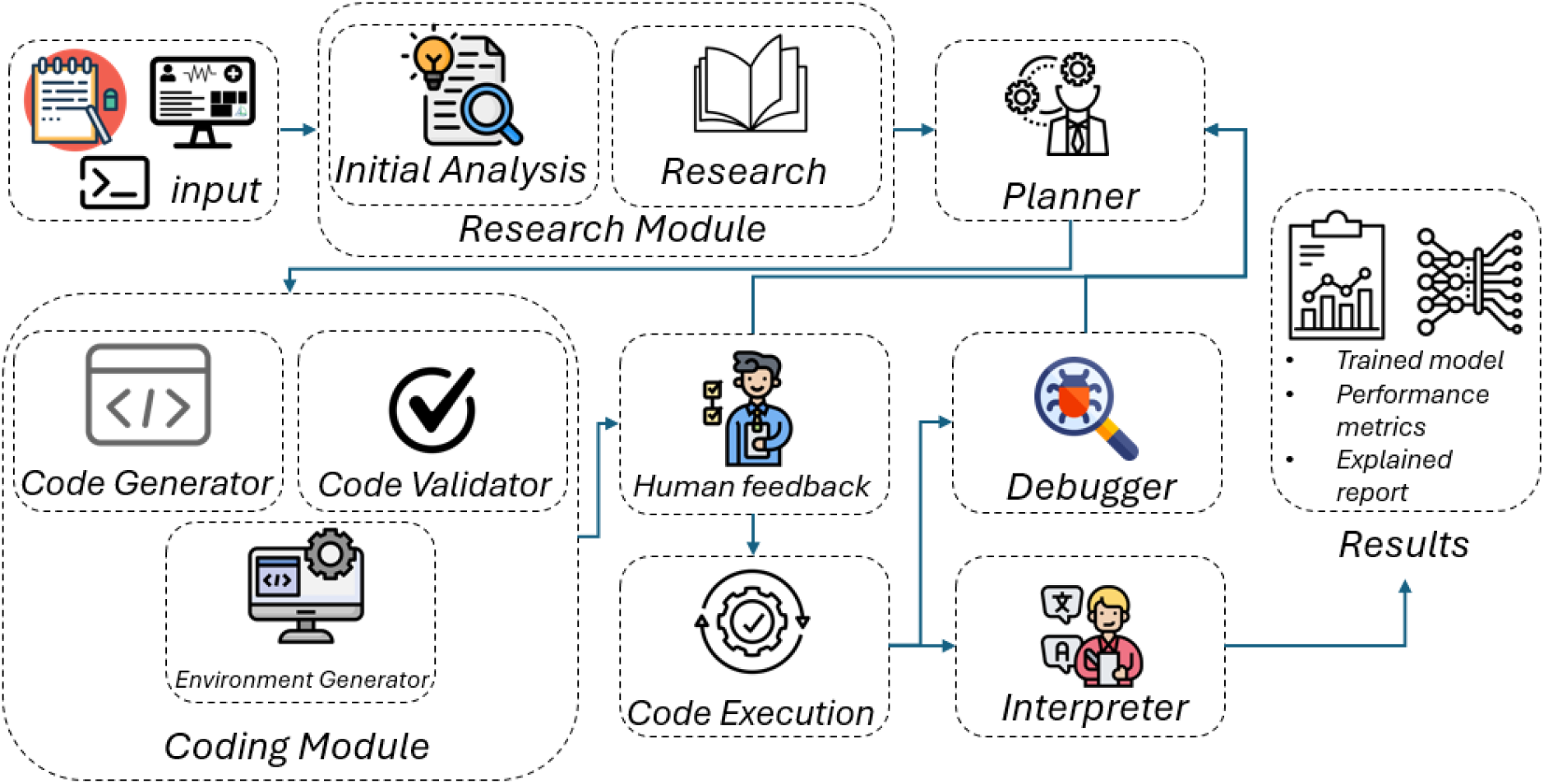
ClinPreAI Agent framework workflow. The autonomous agent system consists of five specialized modules operating in an iterative pipeline: (1) Research Module receives input data (CSV with clinical variables and target labels) and analyzes the problem domain; (2) Planning Module designs a comprehensive machine learning strategy including feature engineering and model selection; (3) Coding Module generates validated code and creates an AWS SageMaker execution environment; (4) Debugging Module automatically identifies and resolves execution errors; (5) Interpretation Module analyzes results and generates comprehensive reports including model performance metrics (F1-score, accuracy, recall, precision), clinical insights, and executable code. User feedback is integrated at multiple stages, enabling iterative refinement until optimal solution quality is achieved. Arrows indicate information flow between modules, with feedback loops shown returning to the Planning Module for strategy adaptation.

**Figure 2.**
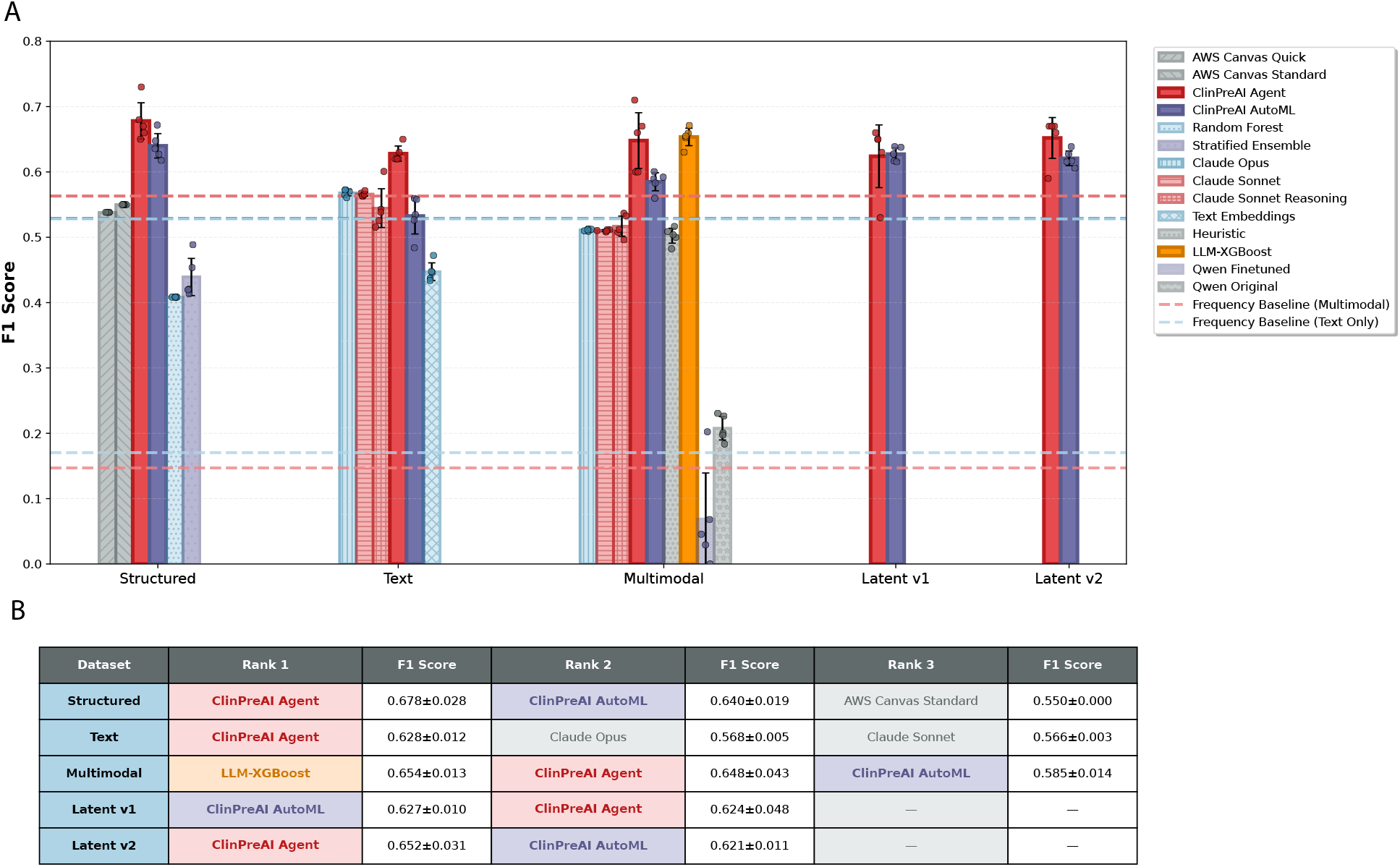
Performance comparison across dataset configurations. **(A)** F1 scores of different methods grouped by dataset type. Bars represent mean F1 scores across 5 different test sets, with error bars indicating standard deviation. Individual data points (dots) show performance in each run. Dashed horizontal lines indicate high majority class baseline performance thresholds (General: 0.563; Text Only: 0.529). ClinPreAI Agent (red) and ClinPreAI AutoML (purple-blue) represent the primary methods evaluated, with LLM-XGBoost (orange) demonstrating specialized performance on multimodal data, while other approaches serve as baselines for comparison. **(B)** Top three performing models for each dataset type, ranked by mean F1 score. The table demonstrates that LLM-XGBoost achieves top performance on multimodal data, ClinPreAI Agent matches top performance on structured data and achieves competitive results across all configurations, while text-only predictions remain challenging across all methods.

**Figure 3.**
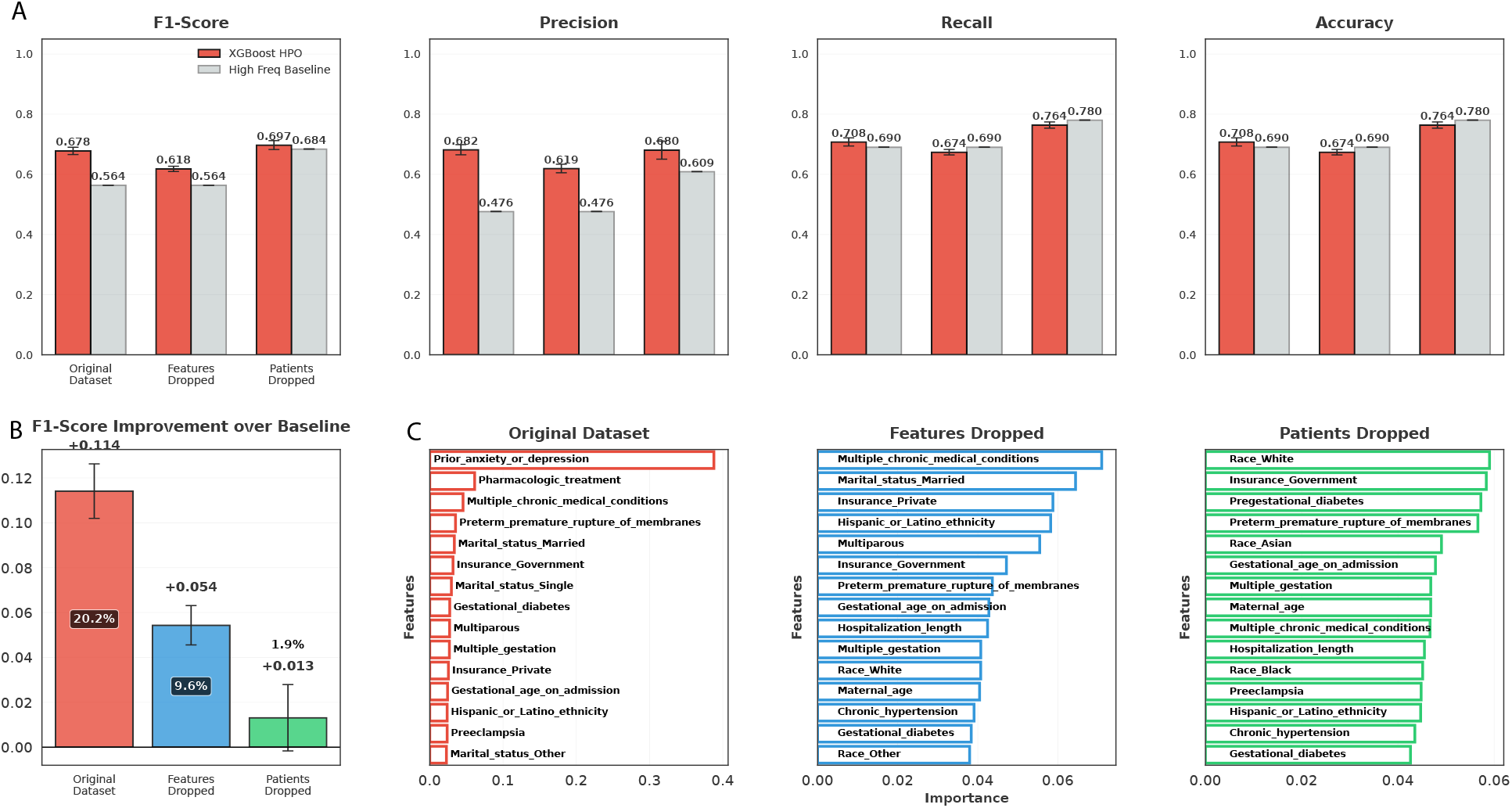
Comprehensive model performance and feature importance analysis across ablation conditions. *Top row (A):* Performance metrics (F1-score, precision, recall, accuracy) comparing hyperparameter-optimized XGBoost (red bars) against high-frequency baseline (gray bars) across three dataset configurations. The original dataset achieved performance (F1=0.68), while the special features dropped configuration showed diminished but non-trivial capability (F1 ≈ 0.62). The patients dropped configuration exhibited near-random performance, with scores indistinguishable from stratified guessing. *Bottom left (B):* XGBoost performance gain over high-frequency baseline reveals a 20% improvement for the original dataset, only 9.6% for the special features dropped dataset, and 1.9% gain for the patients dropped dataset. This demonstrates that while absolute F1 scores may appear higher in the patients dropped condition, this reflects class imbalance rather than genuine predictive power or model capability. *Bottom right (C):* Feature importance analysis across ablation conditions. In the original dataset, prior anxiety or depression history dominates prediction, followed by pharmacological treatment, establishing mental health history as the primary driver. With mental health features removed, the model reallocates importance to chronic medical conditions as a compensatory proxy, though predictive power declines substantially. In the patients dropped configuration, sociodemographic variables (race, insurance status) emerge as nominal top features yet fail to support meaningful discrimination—indicating weak population-level associations rather than individual risk. Only the top 15 features are displayed per condition. Error bars represent standard deviation across five independent experiments with different random seeds. The convergence of all models to baseline performance in the patients dropped condition demonstrates that predictive signal derives primarily from recurrence risk rather than de novo identification.

**Figure 4.**
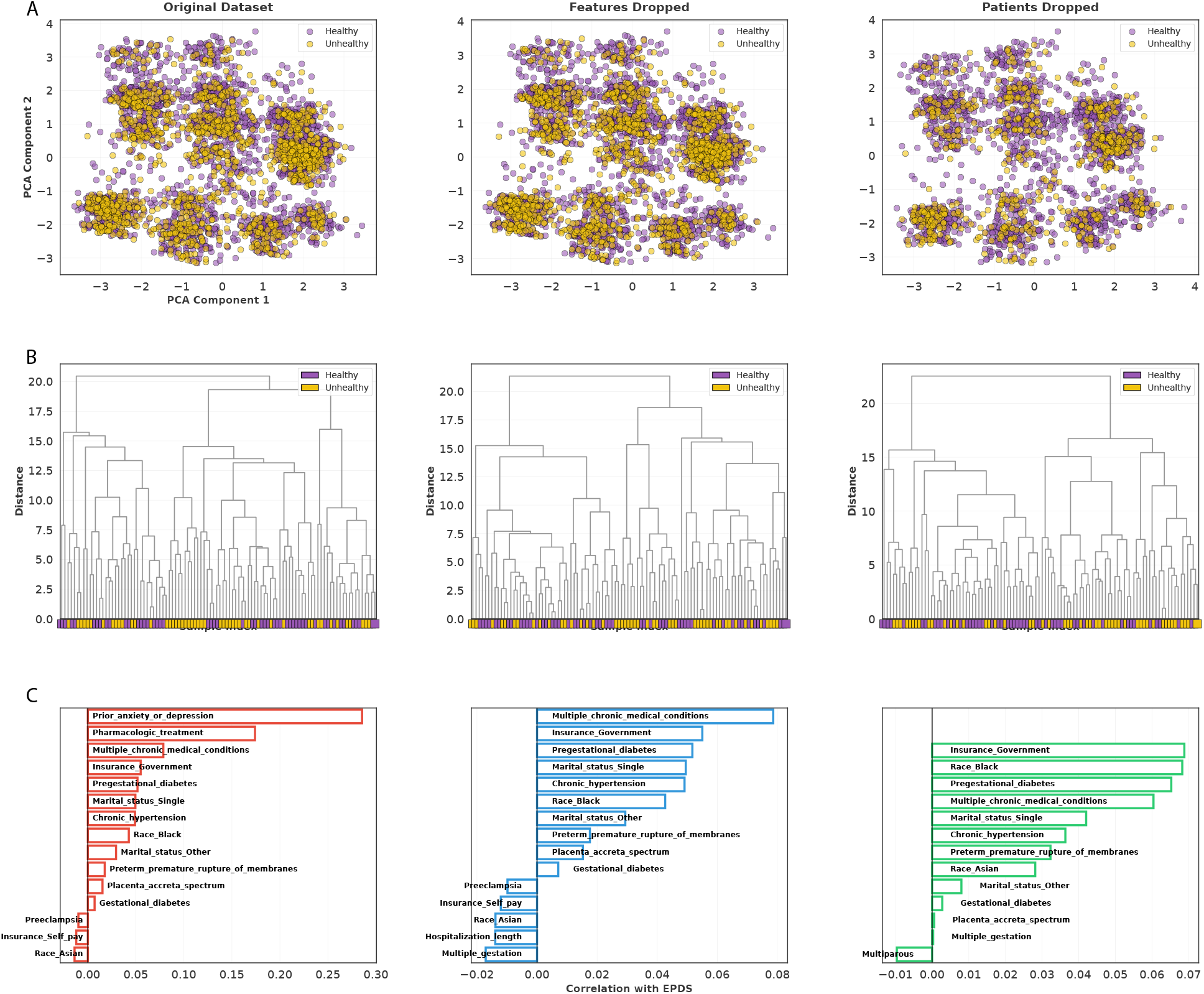
Unsupervised learning analysis reveals lack of natural clustering and identifies key correlates of perinatal depression risk. Unsupervised learning complementary analyses were performed across three dataset configurations: Original Dataset (left column), Features Dropped (middle column, excluding depression history variables), and Patients Dropped (right column, excluding patients with prior depression history). **(A)** Principal Component Analysis (PCA) demonstrates substantial overlap between healthy (purple) and unhealthy (yellow, EPDS ≥ 10) patient groups, with no clear separation in the first two principal components. Other dimensionality reduction visualizations like UMAP and T-SNE yield in similar results. **(B)** Hierarchical clustering dendrograms further confirm the absence of natural patient subgroups. Colored squares at the base represent individual patient EPDS scores, and their intermixing throughout dendrogram branches indicates depression risk is not associated with distinct clustering patterns. Sample size was limited to 100 patients per dendrogram for visualization clarity. **(C)** Feature correlation analysis identifies the top 15 features most strongly associated with EPDS scores in each dataset. In the original dataset, depression history variables show expected strong positive correlations. After removing these features, chronic health conditions and socioeconomic factors emerge as primary correlates. In the patients-without-history subset, demographic and metabolic factors demonstrate the strongest associations with depression risk. The relatively modest correlation coefficients across all features suggest that perinatal depression is a complex, multifactorial condition that cannot be explained by simple linear relationships with individual clinical variables.

**Figure 5.**
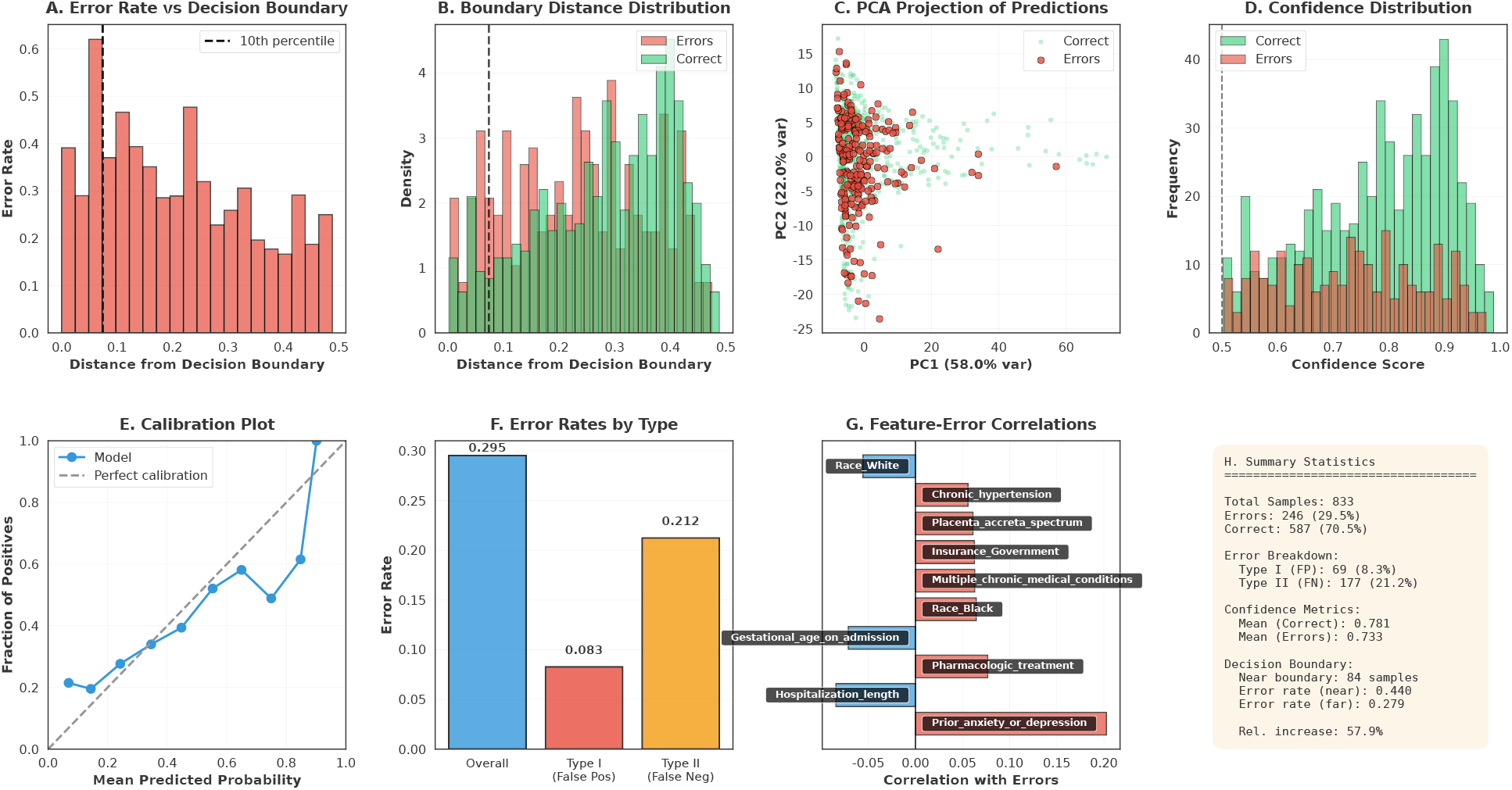
Comprehensive error analysis of XGBoost model performance. (A) Error rate as a function of distance from decision boundary, showing elevated errors near classification threshold with 10th percentile marked. (B) Distribution of boundary distances comparing correct predictions vs. errors, revealing errors concentrate near the decision boundary. (C) PCA projection of test samples with errors highlighted, showing spatial distribution of misclassifications in reduced feature space. (D) Confidence score distribution comparing correct predictions vs. errors, with decision threshold at 0.5. (E) Calibration plot comparing predicted probabilities to observed frequencies, with calibration reference line. (F) Error rates stratified by type, including overall error rate, Type I errors (false positives), and Type II errors (false negatives). (G) Feature-error correlations for top 10 features, showing positive (red) and negative (blue) associations with prediction errors. (H) Summary statistics panel displaying sample counts, error breakdowns, confidence metrics, and decision boundary analysis including relative error rate increases near the boundary.

The agent system operates through the following modules:

#### Research Module

Receives the input CSV file containing structured and unstructured data (one patient per row) along with the target column specification (EPDS high/low label) and optional user prompts. The module enhances its knowledge by analyzing clinical variables and sample data, leveraging its internal knowledge base to understand the problem domain.

#### Planning Module

Receives research insights and designs a comprehensive approach for training a classification model. The planner creates a detailed strategy including feature engineering, model selection, and evaluation metrics tailored to the predictive task.

#### Coding Module

Generates validated code based on the planning module’s specifications. Creates an AWS SageMaker environment for code execution and presents the generated code to the user for approval.

#### Debugging Module

Automatically identifies and resolves execution errors in the approved code, ensuring robust implementation through systematic error detection and correction.

#### Interpretation Module

Analyzes and explains the results, generating comprehensive reports on model performance and clinical insights. Produces trained predictive models, performance metrics (F1-score, accuracy, recall, precision), interpretable reports explaining model decisions and clinical relevance, and executable code for reproducibility.

User feedback on the proposed approach is routed back to the planning agent for redesign. This iterative process continues until user satisfaction is achieved, allowing the system to adapt its strategy based on domain expertise and performance requirements. The system maintains convergence tracking and implements stopping criteria to prevent excessive iteration while ensuring optimal solution quality.

### Baseline and Comparison Models

#### ClinPreAI AutoML Framework

We developed a non-agentic automated machine learning pipeline specifically designed for clinical risk prediction using multimodal healthcare data. The framework integrates structured clinical data with unstructured text through comprehensive preprocessing: numerical features undergo median imputation and z-score normalization; categorical variables use one-hot encoding; and clinical text is processed through tokenization, lemmatization, and stop-word removal while preserving medical terminology using NLTK.

Text features were extracted using dense embeddings from AWS Bedrock’s Cohere model (cohere.embed-english-v3) generating 1,024-dimensional vectors. Model selection employed Bayesian optimization with Optuna to evaluate eight algorithms (XGBoost, Random Forest, Logistic Regression, Gradient Boosting, SVM, Naive Bayes, Extra Trees, AdaBoost) using stratified 3-fold cross-validation with macro F1-score as the objective. Final performance was assessed on a held-out 20% test set.

ClinPreAI AutoML includes a multi-dimensional error analysis framework performing five complementary analyses: (1) feature-wise error pattern analysis; (2) unsupervised error clustering using K-means with PCA visualization; (3) confidence distribution analysis; (4) feature interaction analysis using pairwise correlation and mutual information; (5) decision boundary analysis. Each analysis generates visualizations and the framework produces comprehensive reports with actionable recommendations for model improvement.

#### AWS Canvas ML

We evaluated AWS SageMaker Canvas AutoML in two modes: Quick Mode (4-20 minute runtime) and Standard Mode (up to 4 hours with extended training and comprehensive optimization).

#### Majority Class Classifier

Predicts the most prevalent class (EPDS <10) for all instances, providing a lower bound for model performance given the 69% negative class prevalence.

#### Default Random Forest

Random Forest classifier with default parameters (no hyperparameter optimization), establishing a simple machine learning baseline.

#### Depression History Heuristic

Rule-based baseline predicting positive EPDS outcomes for any patient with prior depression history (either prior anxiety/depression diagnosis or pharmacologic treatment), testing whether sophisticated models improved upon simple clinical heuristics.

#### Stratified Ensemble Model

Trained separate XGBoost classifiers for patients with and without prior anxiety/depression history, with independent hyperparameter optimization via Optuna for each subpopulation. Predictions were generated using the subgroup-specific model corresponding to each patient’s history status, allowing model architectures to specialize for distinct clinical profiles rather than forcing a single model to capture heterogeneous risk patterns.

**Ablation studies additional models**, we additionally evaluated Logistic Regression (L2-regularized with 1,000 maximum iterations and z-score normalization), Random Forest (100 trees with bootstrapped samples), default XGBoost (100 estimators, learning rate 0.3, maximum depth 6), and Optimized XGBoost with Bayesian hyperparameter optimization using Optuna across 100 trials maximizing cross-validation F1 score.

#### Zero-Shot Claude Models

We implemented direct inference using state-of-the-art large language models without finetuning or few-shot learning. We evaluated Claude Opus 4 and Claude Sonnet 4 (both hosted on AWS Bedrock, model versions us.anthropic.claude-opus-4-20250514-v1:0 and claude-sonnet-4-20250929) on the Text Only dataset. For the Multimodal dataset, tabular variables were serialized and the same Claude models were tested. We additionally evaluated Claude Sonnet 4 with extended reasoning capabilities enabled.

#### Cohere Embeddings + Logistic Regression

Text embeddings from AWS Bedrock Cohere (cohere.embed-english-v3) combined with default-parameter logistic regression, establishing a simple baseline.

#### Fine-Tuned Qwen Model

We fine-tuned a Qwen2.5-7B model using QLoRA via the Unsloth Python package on the Multimodal dataset and compared its performance against zero-shot baselines.

#### LLM-Enhanced XGBoost

We developed a hybrid approach combining large language model inference with gradient boosting to leverage both unstructured clinical narratives and structured data. The pipeline first processes each patient’s social worker notes through Claude Sonnet 4 (us.anthropic.claude-sonnet-4-20250514-v1:0 via AWS Bedrock) to generate pseudo-EPDS scores (0-30 scale) based on a detailed scoring rubric encompassing the ten core EPDS domains (anhedonia, anxiety, self-blame, overwhelm, sadness, crying, self-harm ideation, sleep disturbance, fear, and anticipatory pleasure). These LLM-derived scores are then integrated as additional features alongside the original structured clinical variables. The combined feature set undergoes XGBoost classification with Bayesian hyperparameter optimization via Optuna (50 trials, stratified 3-fold cross-validation, TPE sampler) to identify optimal model configurations. This approach enables the model to utilize both the nuanced semantic understanding of clinical narratives captured by the LLM and the predictive power of structured clinical data, creating a multimodal prediction framework that outperforms traditional feature extraction methods.

### Performance Metrics

Model performance was evaluated using a comprehensive suite of metrics: accuracy (overall correct classification rate), precision (positive predictive value, weighted average), recall (sensitivity or true positive rate, weighted average), F1 score (harmonic mean of precision and recall, weighted average), and ROC-AUC (area under the receiver operating characteristic curve). Weighted averaging accounted for class imbalance, ensuring metrics reflected performance across both positive and negative EPDS outcomes.

For primary experiments, final model performance was assessed on held-out test sets comprising 20% of the data using stratified splits. For all analyses, each model was evaluated using five different random states for train-test splitting with 80/20 stratified partitioning. This multi-seed approach mitigated the influence of data partition variability and enabled calculation of confidence intervals. Results were reported as mean ± standard deviation across all runs.

### Ablation Studies

To systematically evaluate the contribution of depression-related features and patient subpopulations to model performance, we conducted comprehensive ablation studies under three experimental conditions:

#### Complete Dataset

The Structured Only dataset included all 27 clinical features and all 4,161 patients, serving as the performance baseline. This dataset contained two depression-related features: `Prior_anxiety_or_depression` (binary) and `Pharmacologic_treatment` (binary), representing documented mental health history and prior psychotropic medication use.

#### Features-Dropped Dataset

We created a modified dataset by removing both depression-related features while retaining all patients. This configuration tested whether models could accurately predict EPDS outcomes using only non-psychiatric clinical indicators, simulating scenarios where mental health history might be unavailable, unreliable, or intentionally excluded to avoid stigmatization bias.

#### Patients-Dropped Dataset

We excluded all patients with documented depression history (`Prior_anxiety_or_depression = 1` OR `Pharmacologic_treatment = 1`). This subset evaluated model performance on a potentially lower-risk population and tested whether predictive patterns learned from the general population generalized to patients without prior mental health concerns.

Confusion matrices were generated for each model-condition combination to reveal specific misclassification patterns. For tree-based models, we analyzed how feature importance rankings changed across ablation conditions using Spearman rank correlation coefficients, identifying which clinical variables compensated for removed depression-related features.

#### Prior–Mental-Health–History–Only Dataset

For completeness, we conducted a complementary analysis restricted to patients with a documented history of anxiety and/or depression (`Prior_anxiety_or_depression`=1 or `Pharmacologic_treatment`=1). This high-risk subgroup analysis assesses whether model performance differ when prior mentalhealth history is always present. A Venn diagram illustrating the overlap between this subgroup and EPDS-positive cases (as defined in this study), together with F1-score performance plots, is provided in the Supplementary Material.

### Interpretability Analysis

#### Feature Importance Analysis

We performed SHAP (SHapley Additive exPlanations) analysis on best-performing structured-data model from the agent planning, XGBoost, to identify key predictive features and quantify their contribution to model predictions. SHAP values provided both global feature importance rankings and patient-level explanations for individual predictions. This is shown in the Supplemental Material.

#### Error Pattern Analysis

ClinPreAI AutoML’s comprehensive error analysis framework characterized model failure modes through five complementary analyses: (1) feature-wise error correlation to identify variables most strongly associated with misclassifications; (2) unsupervised error clustering using K-means with PCA visualization to discover common patterns among failures, distinguishing Type I and Type II errors; (3) confidence distribution analysis examining model calibration; (4) feature interaction analysis using pairwise correlation and mutual information; and (5) decision boundary analysis quantifying error rates near classification thresholds. We used these figures to manually explore specific cases where the model was misclassifying to understand the nature of these mistakes.

#### Unsupervised Learning Validation

To understand inherent data structure independent of EPDS outcomes, we conducted comprehensive clustering analyses across the three ablation dataset configurations. We applied K-means, DBSCAN, and hierarchical agglomerative clustering, evaluating results using internal validation metrics (Silhouette score, Davies-Bouldin Index, Calinski-Harabasz Index) and external validation against EPDS labels using contingency analysis, chi-square tests, and cluster purity metrics. Correlation analysis identified features most strongly associated with depression risk, and hierarchical feature clustering revealed multicollinear variable groups and latent clinical dimensions.

### Manual error inspection

We randomly sampled 35 model errors and, with mental-health experts, reviewed the associated social-worker notes and case histories to analyze discrepancies between the ground-truth EPDS and the LLM-Enhanced XGBoost pseudo-EPDS prediction and rationale.

### Implementation

All analyses were performed using Python 3.x with scikit-learn (v1.x) for machine learning algorithms and clustering; Optuna for Bayesian hyperparameter optimization; umap-learn for UMAP implementation; scipy for hierarchical clustering and statistical tests; NLTK for text preprocessing; numpy/pandas for numerical computations and data manipulation; and matplotlib/seaborn for statistical visualizations. AWS Bedrock provided access to Claude models and Cohere embeddings. AWS SageMaker hosted the ClinPreAI Agent execution environment. Agent was designed using langgraph. Random states were fixed throughout for reproducibility, with results validated across multiple initializations for stochastic algorithms.

## Data Availability

The data produced in this study are protected under the Institutional Review Board (IRB) regulations of Baylor College of Medicine and are not publicly available. Data can be requested and provided upon approval from the IRB.

## Acknowledgments

This research was supported by a fellowship from the Gulf Coast Consortia on the NLM Training Program in Biomedical Informatics and Data Science (T15 LM007093), by the National Science Foundation Graduate Research Fellowship Program (NSF GRFP Fellow ID 2024370642), by NICHD grant no. 5K12HD103087 (PI M.A. Belfort), and by the Chao Endowment and the Huffington Foundation. This work was partially supported by the use of facilities and resources at the Houston VA HSR&D Center for Innovations in Quality, Effectiveness and Safety (Cin13-413) and the South Central Mental Illness Research, Education, and Clinical Center. The opinions expressed are those of the authors and not necessarily those of the Department of Veterans Affairs, the U. S. Government, or Baylor College of Medicine.

## Author Contributions

**Daniel Palacios** conceived and designed the study, developed the ClinPreAI agent and AutoML frameworks, performed all data preprocessing and feature engineering, conducted all model training and benchmarking experiments, performed statistical analyses and error inspections, generated all figures and tables, and wrote the original manuscript draft.

**Sukru Aras** designed and validated the HPO-based risk scoring methodology, engineered domain-specific features for the Latent v2 dataset, revised statistical testing procedures.

**Yi Zhong** independently conducted a parallel study using BERT-based approaches and contrastive learning methods. Comparative discussions with him provided valuable methodological insights that informed the experimental design and analysis approaches in this work.

**Jeff Zhao** architected and implemented the ClinPreAI agent framework within the AWS cloud infrastructure, developed the multi-module agentic system using LangGraph, and provided technical support for AWS Bedrock integration and SageMaker deployment.

**Sasidhar Pasupuleti** provided computational infrastructure support, managed data security protocols and HIPAA compliance, and facilitated access to protected health information systems.

**Hyun-Hwan Jeong** provided early feedback on the presentation and manuscript, which greatly contributed to refining the study’s direction.

**Emily S. Miller, Terri L. Fletcher, and Alison N. Goulding** provided clinical expertise in perinatal mental health and psychiatry, contributed to study conceptualization and clinical interpretation of findings, conducted independent clinical validation of model predictions, and ensured clinical relevance and real-world applicability of the research.

**Hu Chen** served as senior scientific mentor, provided guidance on study design and research direction, facilitated exploration of novel analytical approaches, contributed to the interpretation of results, and provided critical feedback on multiple manuscript drafts.

**Zhandong Liu** supervised the project as Principal Investigator, secured funding, provided strategic guidance on research objectives.

