## Supplemental File 1 for "CLINPREAI: AN AGENTIC AI SYSTEM FOR EARLY POSTPARTUM DEPRESSION RISK PREDICTION FROM MULTIMODAL EHR DATA"

### 1 Supplementary Material

#### 1.1 Unsupervised Clustering Analysis Supporting Figures

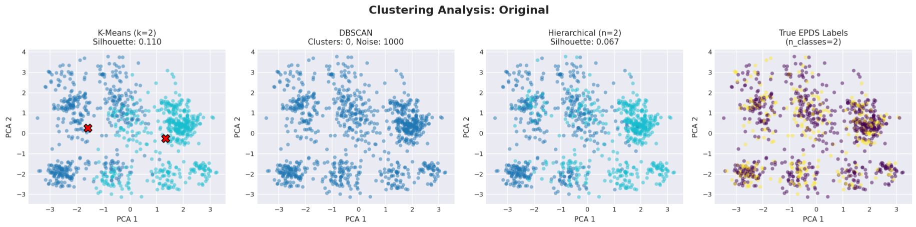


Figure 1: Clustering analysis results for the original dataset, showing patient groupings and cluster characteristics.

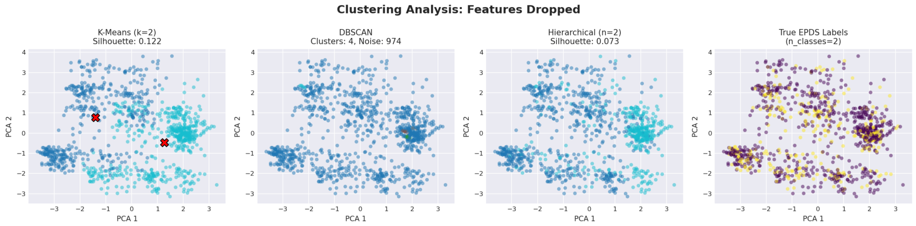


Figure 2: Clustering analysis results for the features dropped dataset.

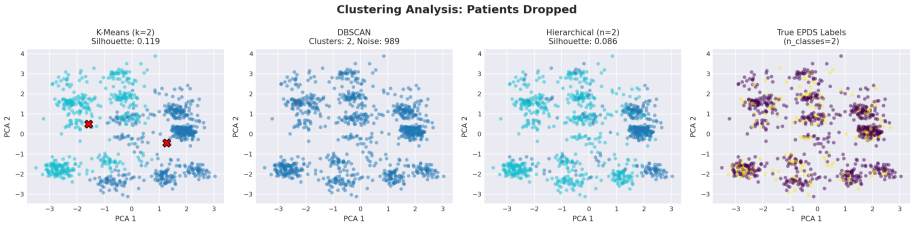


Figure 3: Clustering analysis results for the patients dropped dataset.

**Clustering Performance Summary**

| Dataset | Method | Silhouette Score | N Clusters |
|---|---|---|---|
| Original | KMEANS | 0.110 | 2 |
| Original | DBSCAN | -1.000 | 0 |
| Original | HIERARCHICAL | 0.067 | 2 |
| Features Dropped | KMEANS | 0.122 | 2 |
| Features Dropped | DBSCAN | -0.274 | 4 |
| Features Dropped | HIERARCHICAL | 0.073 | 2 |
| Patients Dropped | KMEANS | 0.119 | 2 |
| Patients Dropped | DBSCAN | -0.257 | 2 |
| Patients Dropped | HIERARCHICAL | 0.086 | 2 |

Figure 4: Summary table comparing clustering metrics across all three datasets.

#### 1.2 Ablation Study Supporting Figures

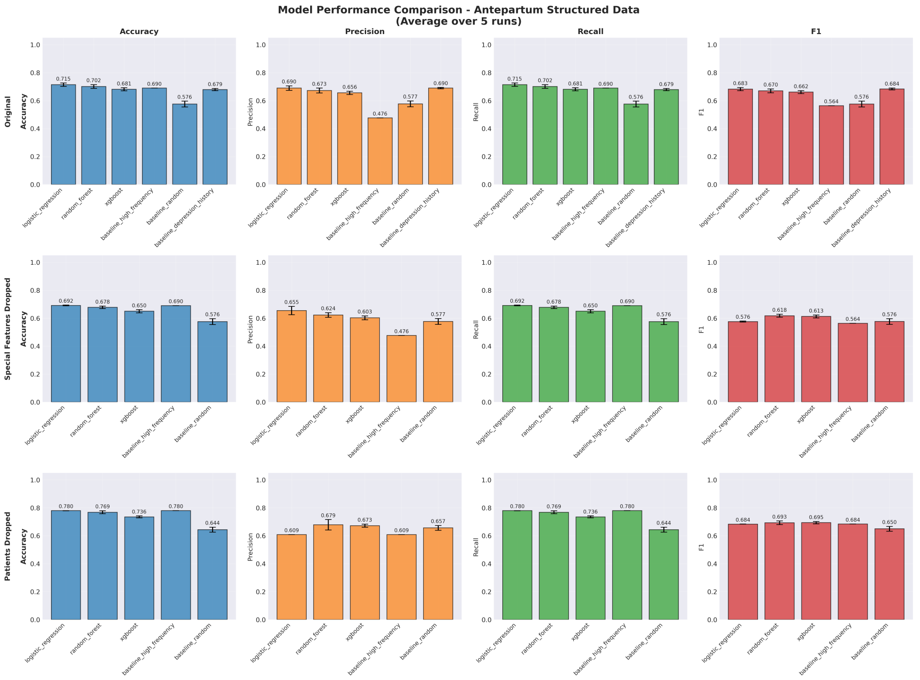


Figure 5: Model performances across all three datasets: Original, Features Dropped, Patients Dropped, from Ablation study. Accuracy, F1 Score, Precision and Recall are being reported for logistic regression, random forest and XGBoost without any hyper-parameter optimization.

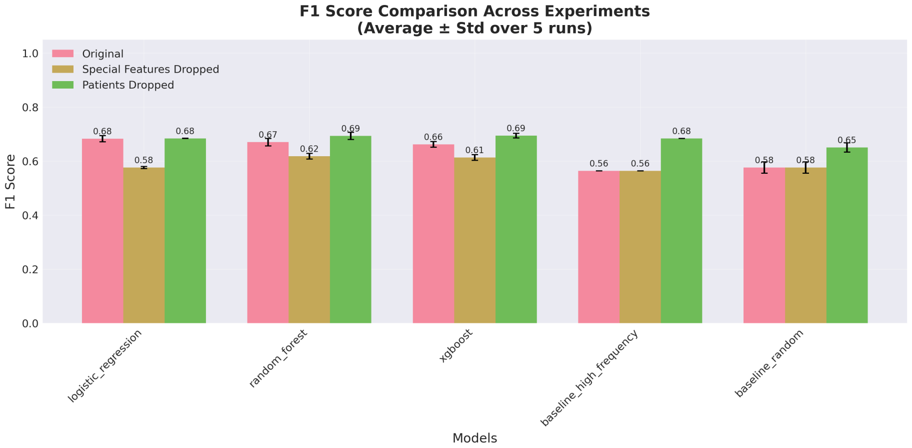


Figure 6: F1 score performances across all three models and baselines for the three datasets.

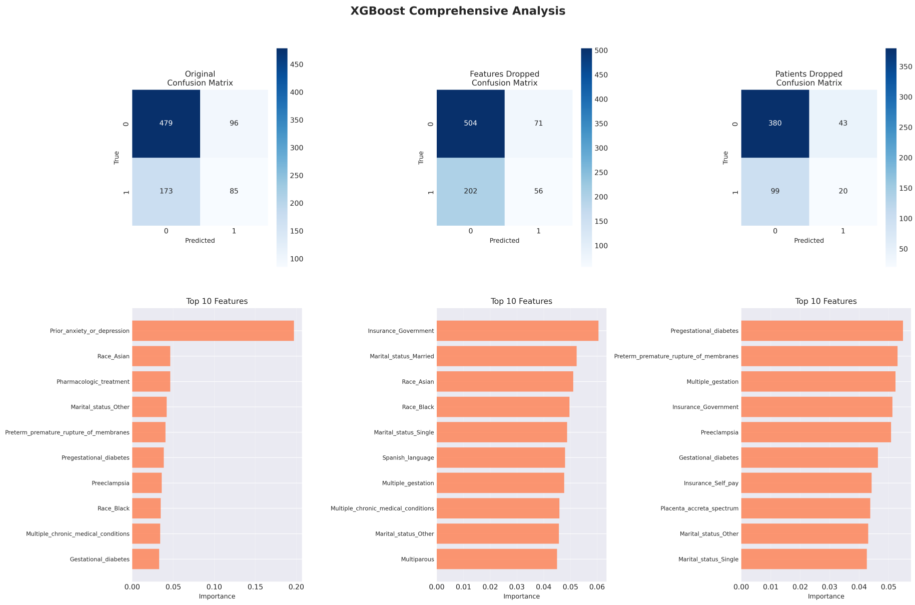


Figure 7: XGBoost (non-optimized) confusion matrices and feature importance for all three datasets.

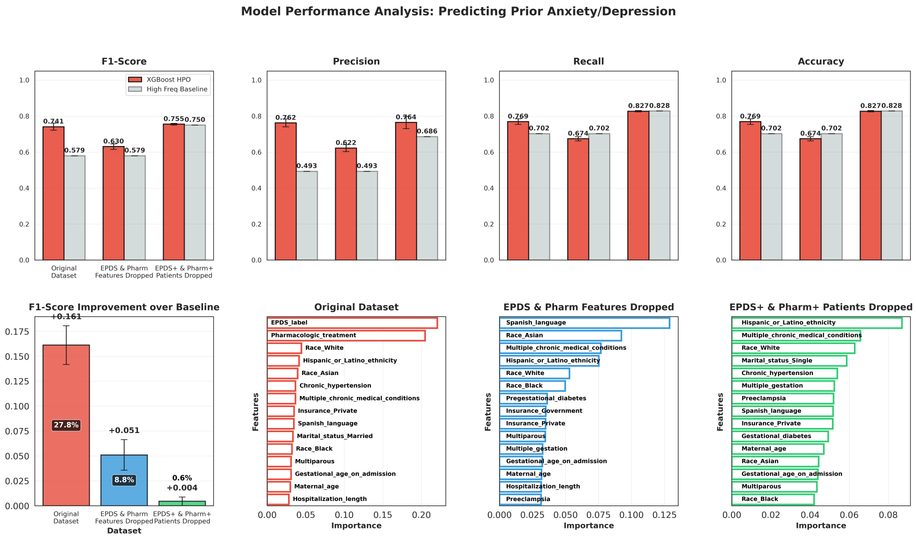


Figure 8: Comprehensive analysis of model performance for predicting outcomes in patients with prior history of anxiety or depression. Eight-panel figure showing performance metrics, confusion matrices, and feature importance across different models.

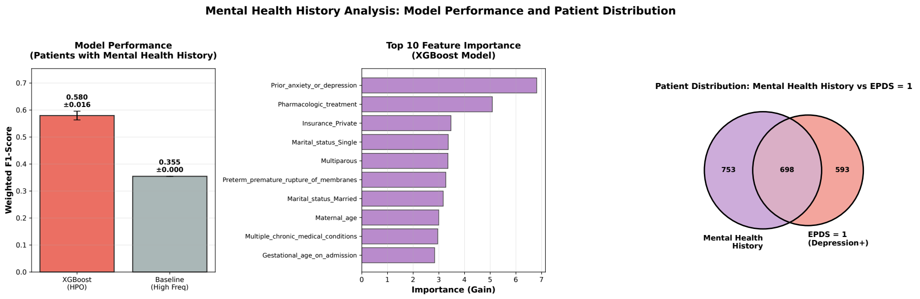


Figure 9: Three-panel analysis of mental health predictions restricted to patients with prior anxiety or depression diagnosis, showing model performance, feature effects, and clinical characteristics.

##### 1.3 SHAP Analysis Supporting Figures

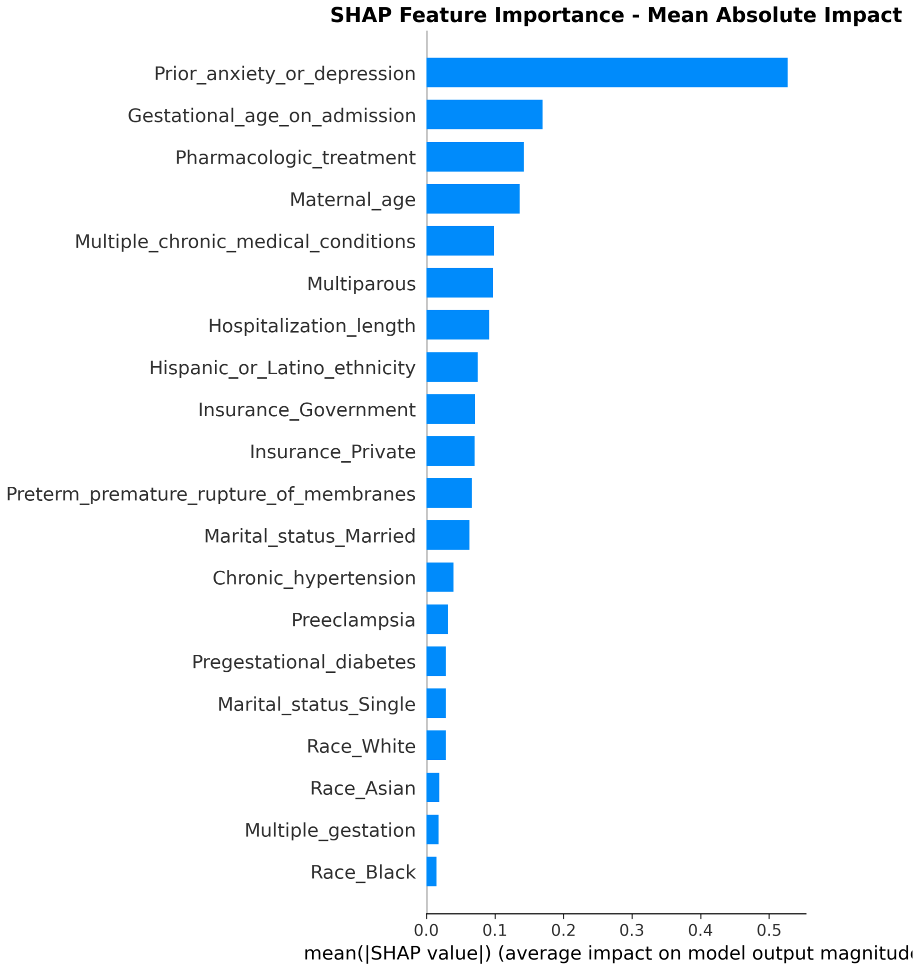


Figure 10: SHAP feature importance bar plot showing the mean absolute SHAP values for each feature, ranked by importance.

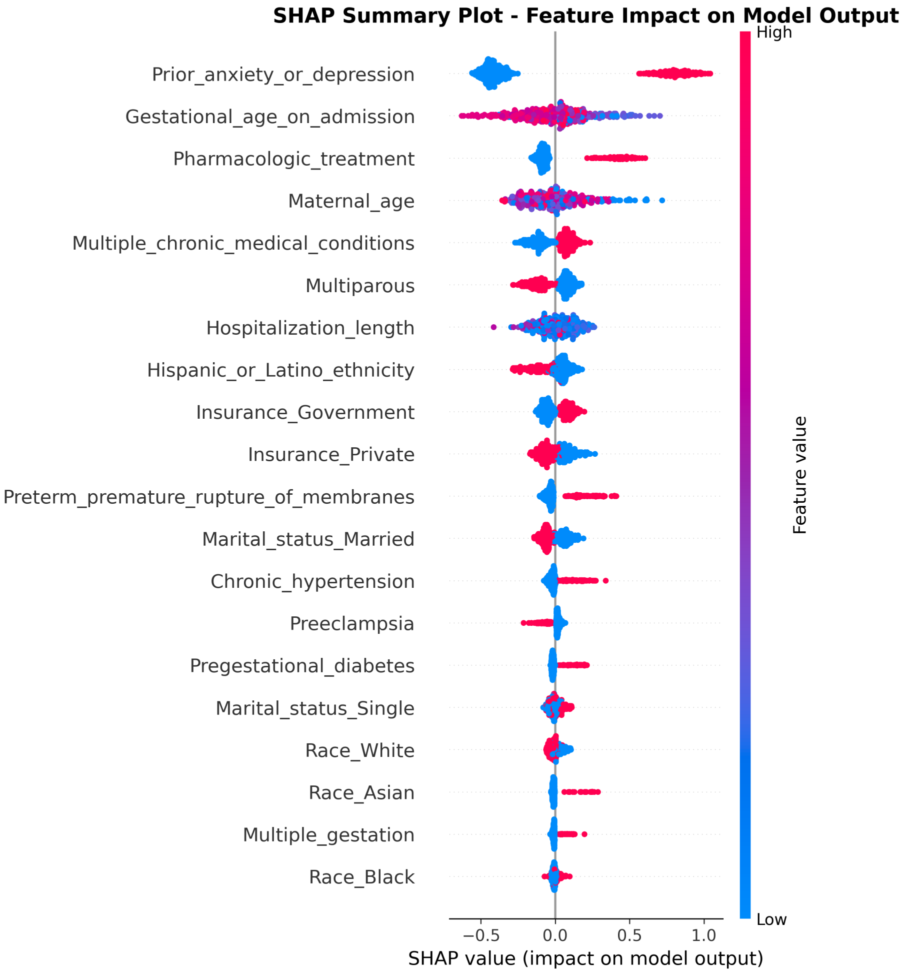


Figure 11: SHAP summary plot displaying the distribution of SHAP values across all features. Each point represents a sample, with color indicating feature value (red = high, blue = low).

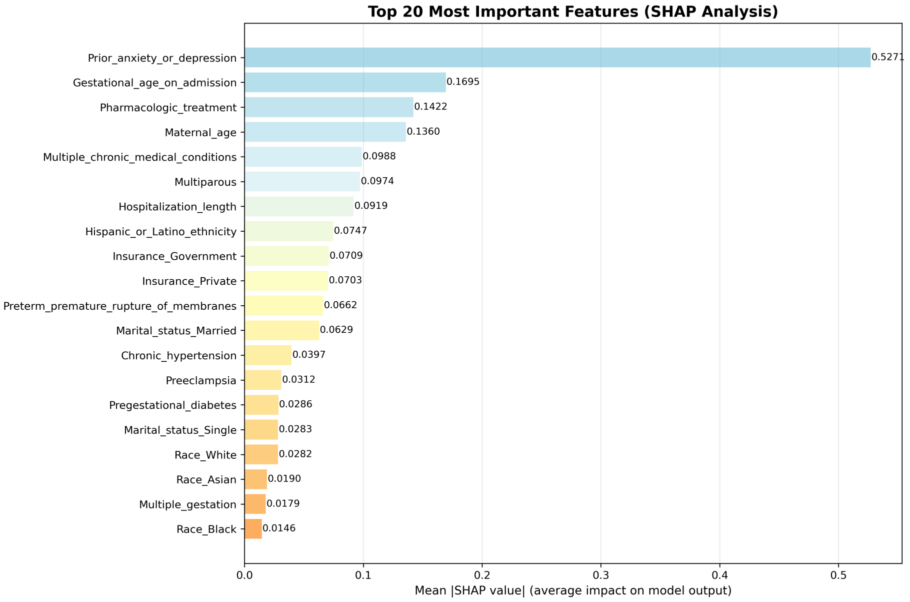


Figure 12: Ranking of top features based on SHAP values.

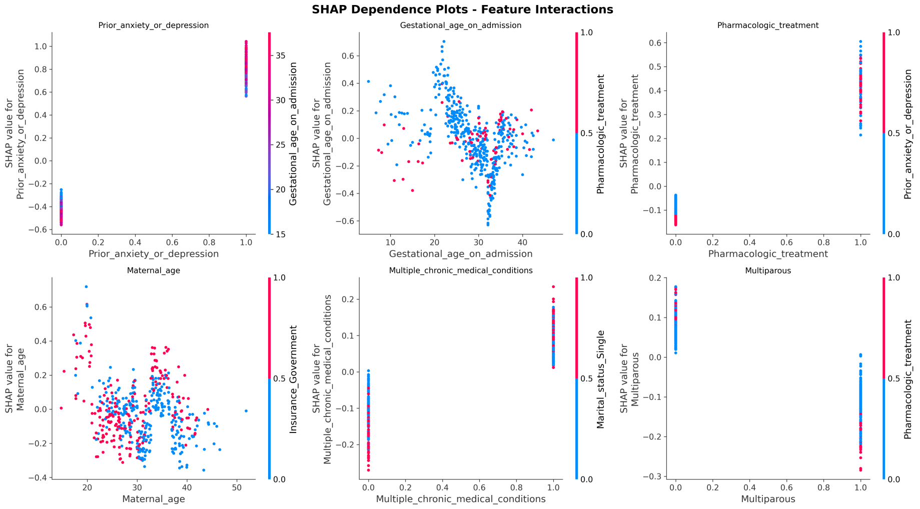


Figure 13: SHAP dependence plots showing the relationship between feature values and SHAP values for key predictive features.

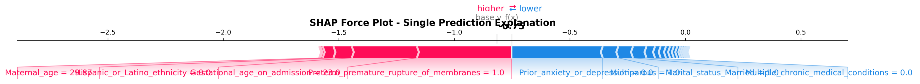


Figure 14: SHAP force plot illustrating how individual features contribute to pushing the model output from the base value to the final prediction.

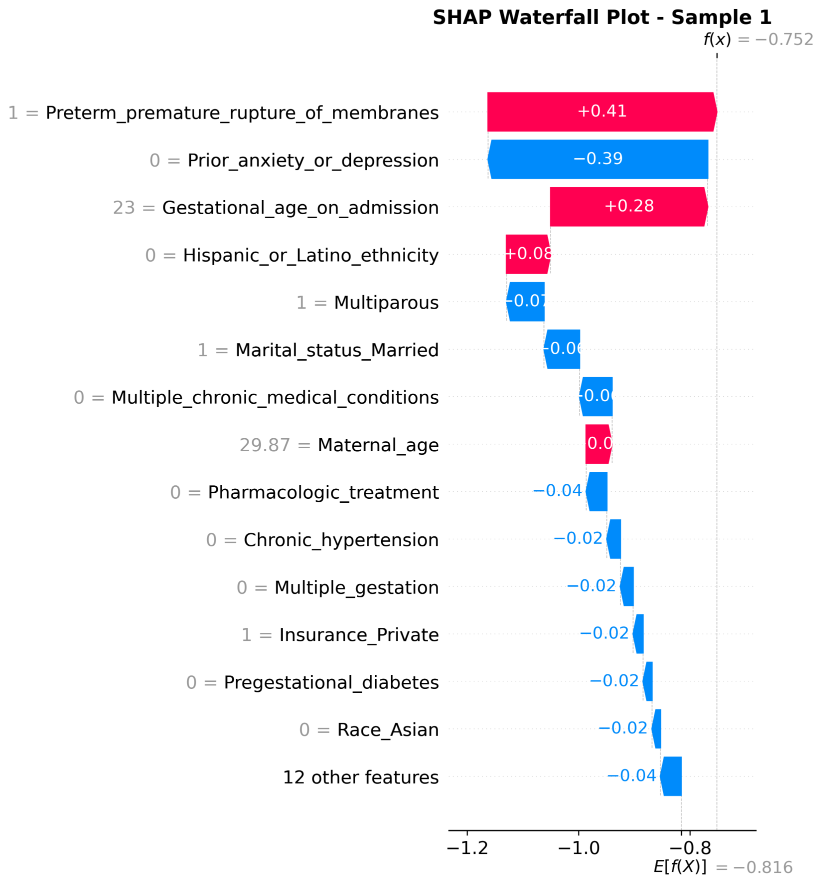


Figure 15: SHAP waterfall plot for sample patient 1, showing the contribution of each feature to the final prediction.

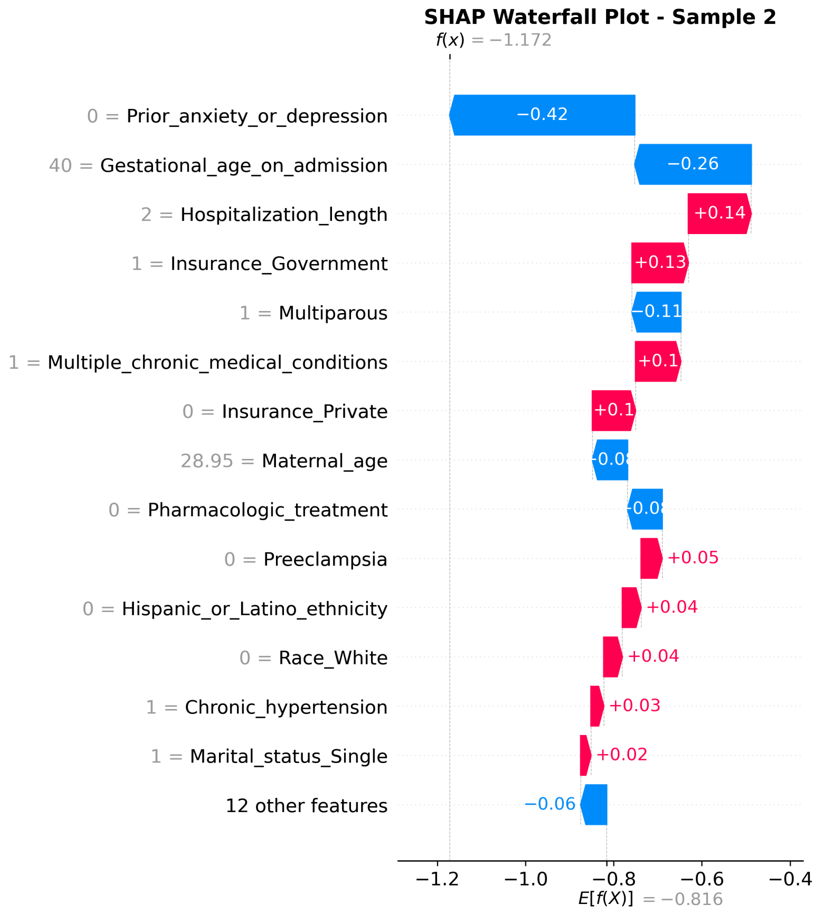


Figure 16: SHAP waterfall plot for sample patient 2, showing the contribution of each feature to the final prediction.

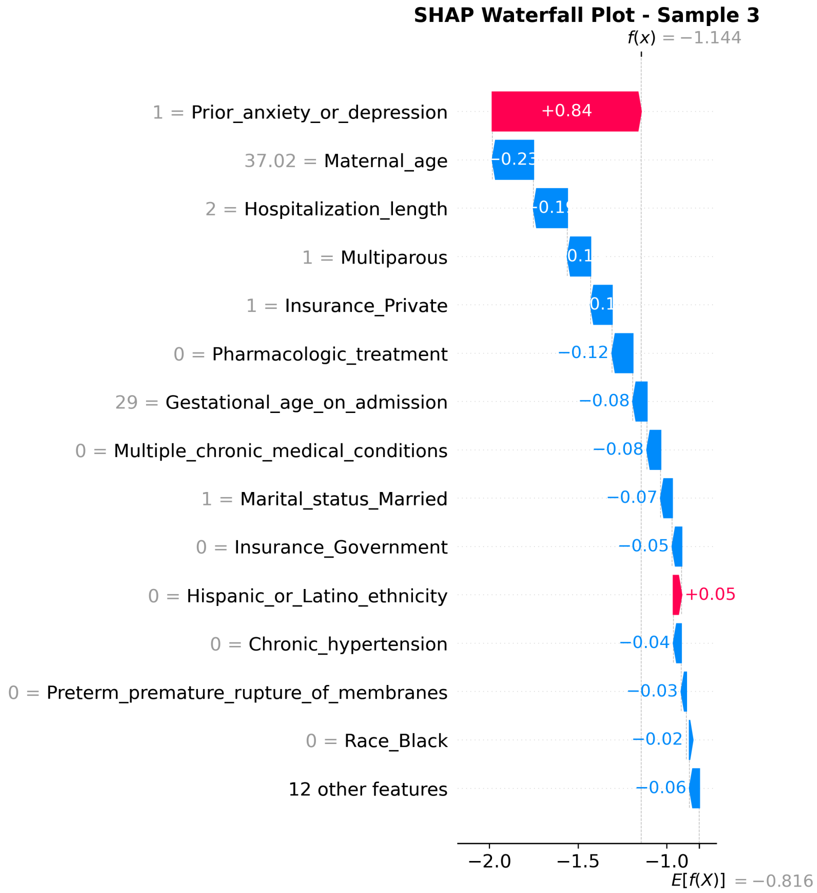


Figure 17: SHAP waterfall plot for sample patient 3, showing the contribution of each feature to the final prediction.

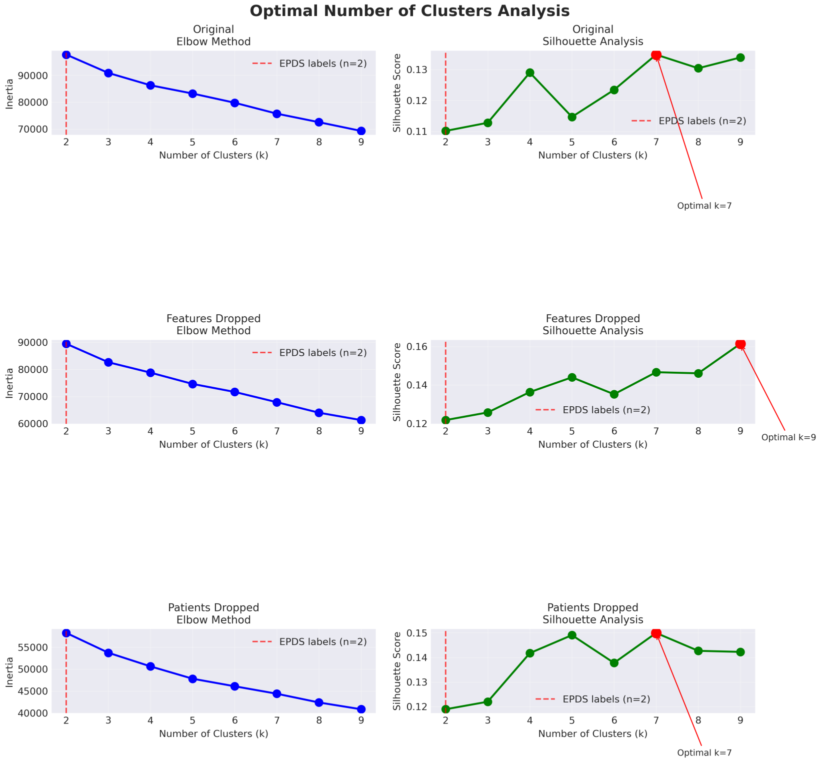


Figure 18: **Optimal cluster determination using elbow method and silhouette analysis.** Left panel shows the elbow plot of within-cluster sum of squares (WCSS) as a function of cluster number, with the elbow occurring around k=7-9 clusters, indicating diminishing returns in explained variance beyond this point. Right panel displays silhouette scores across different cluster numbers, with maximum scores around 0.15 achieved at k=7-9, suggesting weak but optimal cluster cohesion at this configuration. The low silhouette scores across all k values indicate that patients do not form well-separated, discrete subgroups, supporting the interpretation that postpartum depression risk exists along a continuum rather than in distinct categories.
